# How much should we sequence? An analysis of the Swiss SARS-CoV-2 surveillance effort

**DOI:** 10.1101/2023.08.28.23294715

**Authors:** Fanny Wegner, Blanca Cabrera-Gil, Tanguy Araud, Christiane Beckmann, Niko Beerenwinkel, Claire Bertelli, Matteo Carrara, Lorenzo Cerutti, Chaoran Chen, Samuel Cordey, Alexis Dumoulin, Louis du Plessis, Marc Friedli, Yannick Gerth, Gilbert Greub, Adrian Härri, Hans Hirsch, Cedric Howald, Michael Huber, Alexander Imhof, Laurent Kaiser, Verena Kufner, Stephen L. Leib, Karoline Leuzinger, Etleva Lleshi, Gladys Martinetti Lucchini, Mirjam Mäusezahl, Milo Moraz, Richard Neher, Oliver Nolte, Alban Ramette, Maurice Redondo, Lorenz Risch, Lionel Rohner, Tim Roloff, Pascal Schläpfer, Katrin Schneider, Franziska Singer, Valeria Spina, Tanja Stadler, Erik Studer, Ivan Topolsky, Alexandra Trkola, Daniel Walther, Nadia Wohlwend, Cinzia Zehnder, Aitana Neves, Adrian Egli, the SPSP consortium

## Abstract

**Background:** During the SARS-CoV-2 pandemic, many countries directed substantial resources towards genomic surveillance to detect and track viral variants. There is a debate over how much sequencing effort is necessary in national surveillance programs for SARS-CoV-2 and future pandemic threats.

**Aim:** We aimed to investigate the effect of reduced sequencing on surveillance outcomes in a large genomic dataset from Switzerland, comprising more than 143k sequences.

**Methods:** We employed a uniform downsampling strategy using 100 iterations each to investigate the effects of fewer available sequences on the surveillance outcomes: (i) first detection of variants of concern (VOCs), (ii) speed of introduction of VOCs, (iii) diversity of lineages, (iv) first cluster detection of VOCs, (v) density of active clusters, and (vi) geographic spread of clusters.

**Results:** The impact of downsampling on VOC detection is disparate for the three VOC lineages, but many outcomes including introduction and cluster detection could be recapitulated even with only 35% of the original sequencing effort. The effect on the observed speed of introduction and first detection of clusters was more sensitive to reduced sequencing effort for some VOCs, in particular Omicron and Delta, respectively.

**Conclusion:** A genomic surveillance program needs a balance between societal benefits and costs. While the overall national dynamics of the pandemic could be recapitulated by a reduced sequencing effort, the effect is strongly lineage dependent – something that is unknown at the time of sequencing – and comes at the cost of accuracy, in particular for tracking the emergence of potential VOCs.

## Introduction

The emergence of SARS-CoV-2 rapidly became one of the most challenging public health situations for national health agencies around the world. Initially, diagnostic capacities were limited and had to be established. As it became clear that the pandemic was not going to be contained quickly, the evolution of the virus became an important focus of attention as new variants with different worrisome properties emerged, such as higher transmission rates or reduced vaccine effectiveness. Diagnostic assays had to be adapted to distinguish these new emerging viral variants. Classic epidemiology based on incidence data does not provide sufficient resolution and information to direct public health policies in the face of an ever-changing virus [1]. Genome sequencing, meanwhile, can help guide responses on multiple fronts: it allows the analysis of pathogen evolution and transmission, the detection of outbreaks, as well as informs the development and optimisation of diagnostic assays and vaccines – all of which in turn inform public health interventions [1–11].

Consequently, significant effort was directed towards capacity building and creating a fast and reliable surveillance system that allowed the tracking and tracing of (novel) SARS-CoV-2 variants. In Switzerland, individual laboratories quickly stepped up to the challenge of genomic surveillance, but without official targets. This changed in March 2021 with the establishment of the national genomic surveillance program by the Federal Office of Public Health (FOPH), which aimed to sequence 2,000-3,000 samples per week. Due to changed circumstances such as the wide availability of vaccines, this target was adjusted in April 2022 to 500 samples per week with a focus on hospitalised patients (**figure S1**). To date, Switzerland has shared around 161k genome sequences with GISAID (https://gisaid.org/submission-tracker-global/, last accessed 09 May 2023) making it 8^th^ country in the world by number of submitted sequences when normalised by population. Nationally, the Swiss Pathogen Surveillance Platform (SPSP) has become the officially mandated Swiss SARS-CoV-2 Data Hub by the FOPH, providing both a national platform as well as sharing data to other databases such as GISAID and ENA [12,13].

The number of sequences influences how early emerging viral lineages can be detected [14]. Using genomic data from SPSP collected between the onset of the pandemic in February 2020 until the beginning of August 2022, we aim to describe the national surveillance effort and explore how the amount of sequencing affects key surveillance outcomes: (i) first detection of variants of concern (VOCs), (ii) speed of introduction of VOCs, (iii) diversity of lineages, (iv) first cluster detection of VOCs, (v) density of active clusters, and (vi) geographic spread of clusters.

## Results

### Genomic surveillance in Switzerland

A total of 143,260 sequences were available for a population of 8.7 million inhabitants over the observed period in our dataset (cf. Methods). The median percentage of cases that were sequenced each week was 7% (**figure 1**). In total, this constitutes 3.6% of all cases (n=3.95 million) for the whole study period.

**Figure 1.**
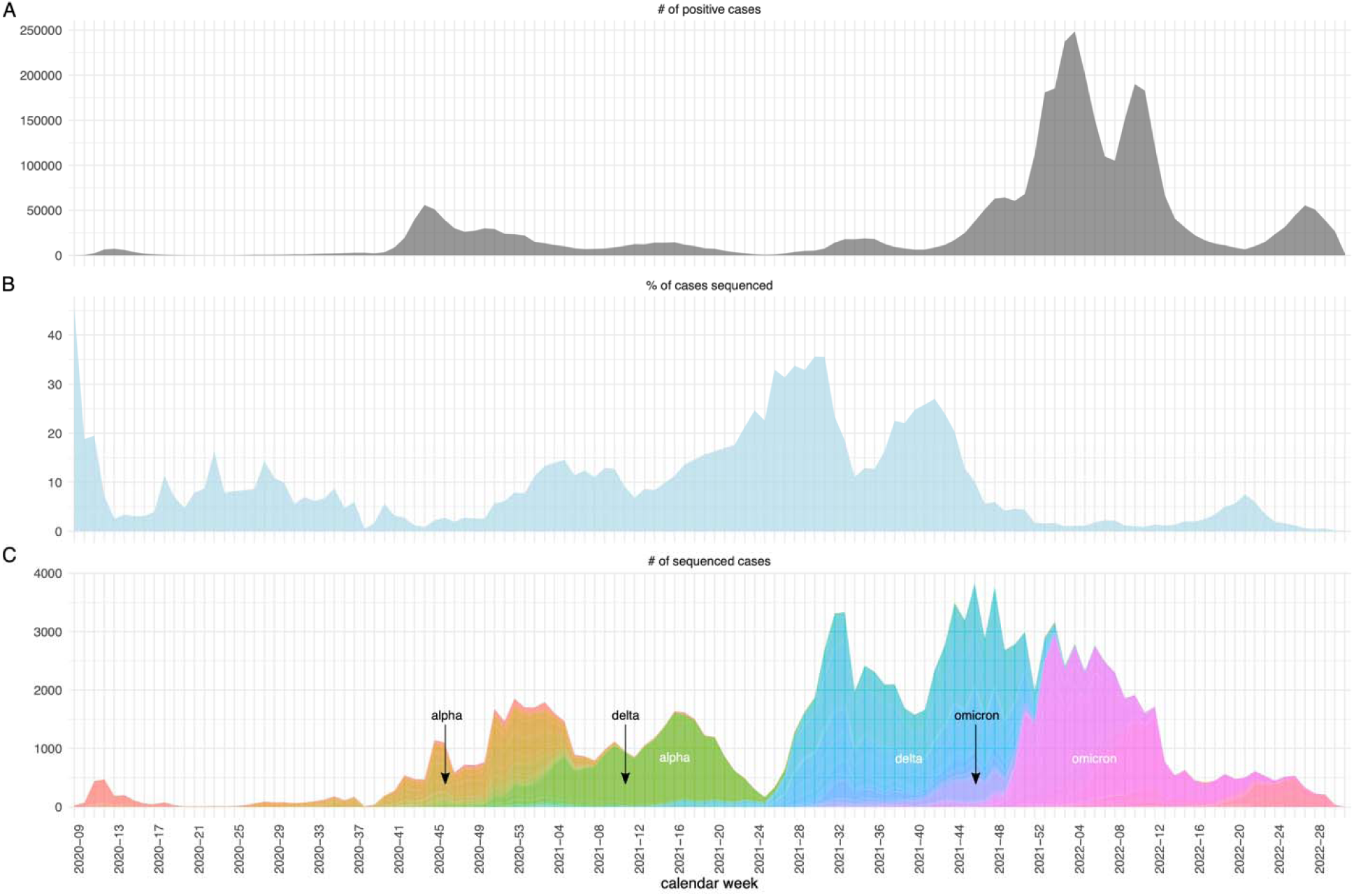
(A) Number of positive COVID-19 cases in Switzerland per week. (B) Percentage of cases that were sequenced per week. C) Number of sequenced cases per week coloured by their Pango lineage assignment. This represents the total number of sequences submitted to SPSP (n=143,260). The black arrows denote the first detection of the VOCs Alpha, Delta, and Omicron. The green wave corresponds to the Alpha wave, the blue and purple colours to the Delta wave, and the pink colours to Omicron.

The first year of the pandemic was characterised by the emergence and persistence of multiple different viral lineages that led to an increase in nucleotide diversity and the lineage diversity index (LDI, as measured by Shannon diversity index, cf. Methods) (**figure 1C** and **figures S2-S6**). This diversity on the nucleotide and lineage level was reduced with the first VOC, Alpha, rising to dominance (**figure S3**). All subsequent waves were dominated by a single VOC, with a slow rise in genetic diversity over time and a peak when a new, usually more divergent VOC was introduced (**figure S3A**). In contrast to Alpha, the Pango nomenclature accommodated for the diversification of Delta and Omicron by introducing many more sublineages. This leads to a LDI when all Pango-designated sublineages are considered, but an overall low LDI when these sublineages are consolidated to VOC labels (**figure S3B**). Moreover, as indicated by the arrows in **figure 1C**, Alpha and Delta were already present in Switzerland several weeks before they increased in frequency and truly emerged as the dominant variant. For Omicron, however, this initiation period was much shorter. The pandemic context during the emergence of each VOC was also very different. For example, case numbers were high at the time of introduction of Alpha and Omicron, but low for Delta (**figure S4**). Delta’s rapid growth phase started briefly after a period of decreasing Alpha cases which coincided with relaxed health and safety measures in Switzerland. This – in addition to its intrinsically higher transmissibility [15] – likely played a role in its rapid increase at the end of June 2021.

### Effect of downsampling on VOCs lineage diversity index

To investigate how the observed LDI would have changed with lower sequencing intensity, we downsampled our dataset (cf. Methods).

Naturally, downsampling had an influence on the absolute number of lineages detected (**figure S6**). However, downsampling had only a minimal effect on the LDI during VOC dominated periods, especially for Alpha and Delta (**figure S5**). The impact of downsampling was stronger during the initial period of the pandemic when the number of available sequences was low. There was also a stronger effect during the later Omicron wave (i.e. once Delta had stopped circulating widely in Switzerland). This might be explained by the fact that Omicron has a highly skewed sublineage distribution with a few widespread sublineages co-circulating alongside an assortment of rare lineages, whereas Delta sublineages are more evenly distributed (**figure 1**), leading to the LDI being more sensitive to the disappearance of rarer sublineages with increased downsampling in Omicron.

### Effect of downsampling on first detection of VOCs and speed of introduction

We also investigated how the first detection of the VOCs would have changed with a reduced sequencing effort, simulated by downsampling our dataset. As expected, the delay of detection increased when fewer sequences were available (**figure 2A**). Surprisingly, downsampling to around a third of the available sequences (50k sequences) only led to a 1 day median delay for Delta and Omicron, and no delay for Alpha. In general, we note that Delta was more sensitive to downsampling than Alpha and Omicron, as indicated by the longer tail of the delays upon repeated downsampling.

**Figure 2.**
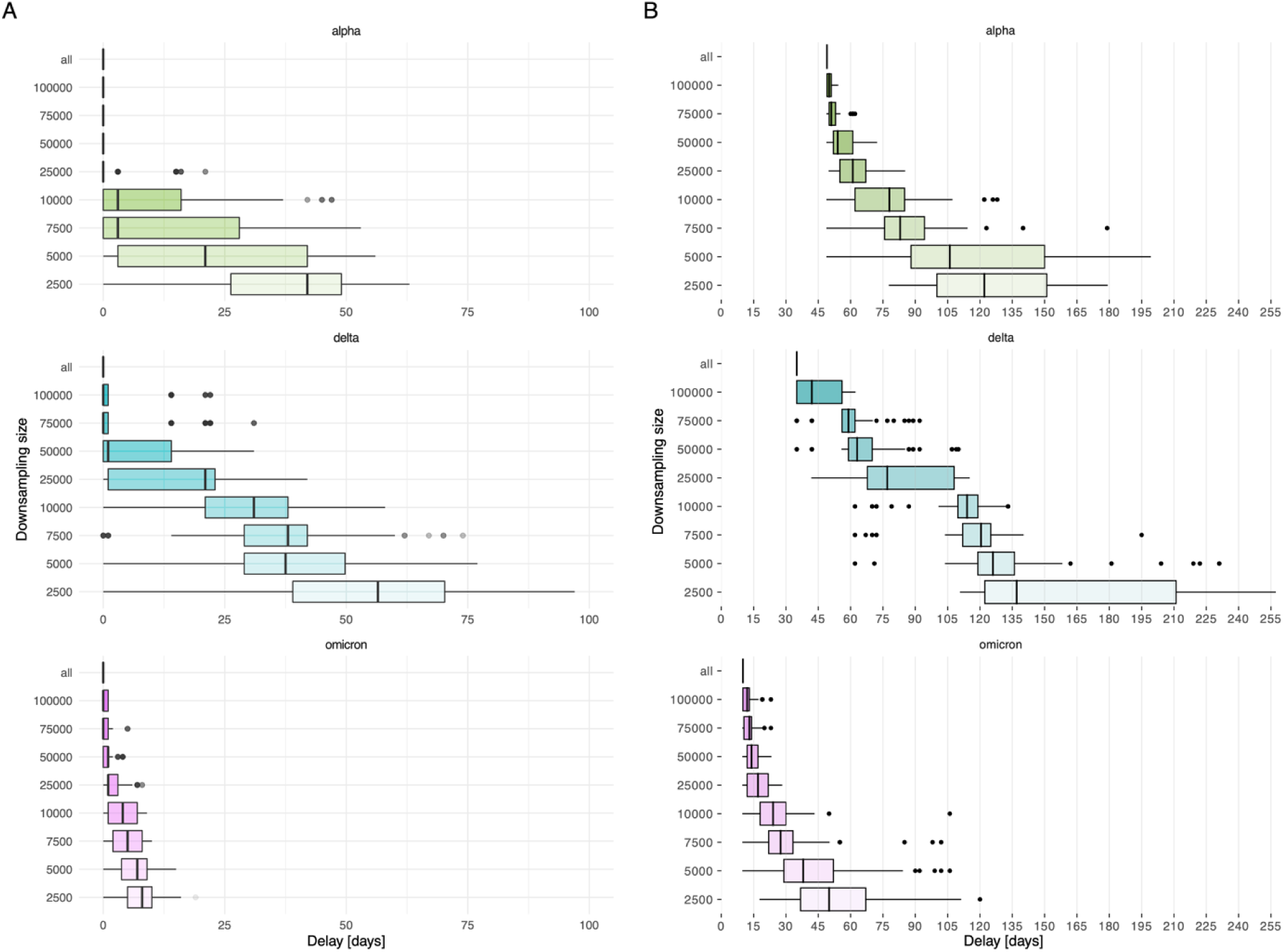
(A) Delay in detecting the first sequence of the VOC upon downsampling, as compared to using the full dataset (“all”). Delays are shown for 100 iterations at each downsampling size. (B) Delay for detecting the first cluster of a VOC after detecting its first introduction, shown for the full dataset (“all”) and upon downsampling. Delays are shown for 100 iterations at each downsampling size. Boxplots display the median and interquartile range, with minimum and maximum values shown with whiskers. Colour code: delays for Alpha in green, for Delta in blue, and for Omicron in pink.

We reasoned that differences in the impact of downsampling might be due to differences in how steeply each VOC rises shortly after introduction. Thus, we further investigated the speed of introduction of each VOC, defined as how quickly a newly introduced VOC was established and grew in frequency. While speed of introduction was similar for Alpha and Delta, it was much faster for Omicron (**figure 3A**). We observed that the speed of introduction also depended strongly on downsampling size, used as a proxy for sequencing effort. A higher sequencing effort was most accurate in capturing the respective growth of the emerging VOCs. This is because strongly downsampled data might not behave in a linear fashion anymore (cf. Methods), leading to arbitrary estimation values driven by sampling bias. Rather than being a fault, this highlights that accurately estimating the speed of growth of a VOC becomes difficult with sparse data. Interestingly, the estimates of the downsampled datasets converge with increased sampling size towards the value calculated for the complete dataset (red line in figure 3), suggesting that the actual sequencing effort captured true VOC dynamics accurately.

**Figure 3.**
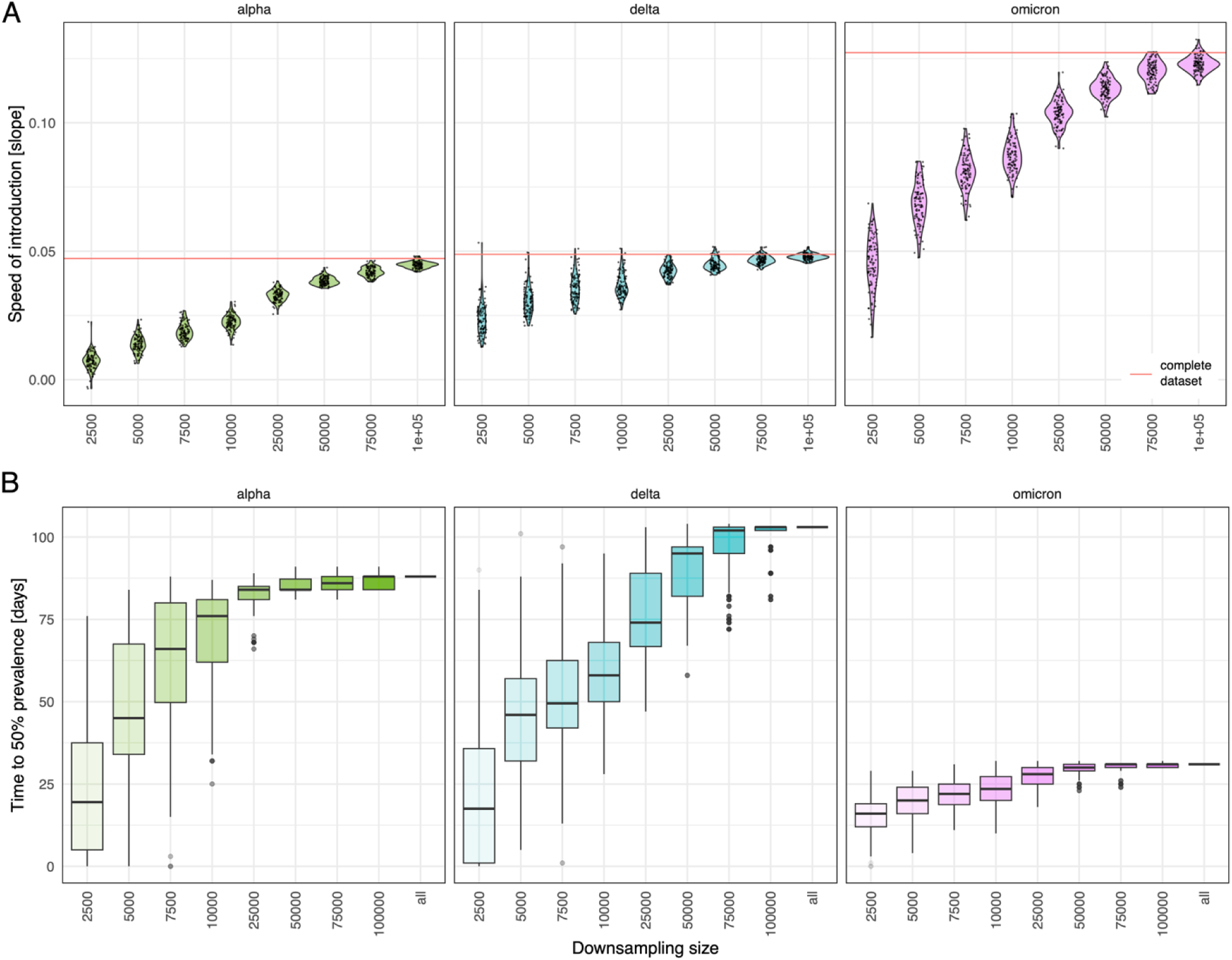
(A) The speed of introduction (i.e. the slope of the linear model of the growth phase of each VOC) upon downsampling in relation to the complete dataset (indicated by the red line). (B) Number of days required to reach 50% prevalence upon downsampling compared to using the full dataset (“all”).

We also examined how long it takes for a VOC to reach dominance or 50% prevalence (**figure 3B**). In the full dataset, Alpha and Delta required 88 days and 103 days, respectively, while Omicron required only 31 days. With increased downsampling, the time needed to reach 50% prevalence decreased. In other words, lineages are seen as dominant earlier when fewer sequences are available. Delta was very sensitive for this measure as the spread of data and also the median time in reaching prevalence increases stronger with each downsampling. However, downsampling to around a third (50k sequences) accelerates prevalence only by 8 days median (4 days for Alpha and 1 day for Omicron), recapitulating it still well.

To summarise, we observe that the impact of downsampling on surveillance outcomes are very VOC dependent (**figure S7**). Downsampling affected Alpha least with regards to the delay in detection and speed of introduction, whereas Delta was affected for both measures. For Omicron, the effect of downsampling on its detection was insignificant, while it was very strong for observing its growth.

These observations mean that our above stated reasoning does not hold true, as Alpha and Delta display a similar speed of introduction for the complete dataset but are differently impacted by downsampling. We can observe, however, that Delta has a unique feature. Its introductory period covers the phase in mid-June 2021 when cases (and sequencing) were very low (**figure 1**) and health and safety measures relaxed giving Delta an additional advantage and leading to its rapid rise. This “gap” might be the reason different surveillance measures are so strongly affected by downsampling for Delta.

### Effect of downsampling on VOC cluster detection

To understand how the transmission information changes when having fewer sequences, we also analysed the effect of downsampling on the detection of clusters.

We first looked at the delay between the detection of the first cluster of each VOC and the first sequence of that VOC. A cluster is defined as a transmission event between 3 or more people (cf. Methods). When considering all available sequences, the delay in cluster detection is very VOC dependent: 49 days for Alpha, 35 days for Delta and 10 days for Omicron (**figure 2B**). **Figure 2B** shows the effect of downsampling on the delay to detect clusters. A commonly asked question during the pandemic was whether new cases were due to endemic transmission or new introductions. This is of epidemiological interest as it can affect the backward and forward tracing strategy. Delay upon downsampling was again more pronounced for Delta. When downsampling to 50k sequences, the delay in the detection of the first cluster is much shorter for Alpha (54 days) and Omicron (14 days) but more accentuated for Delta (63 days). The first Delta clusters appeared in May 2021, but it was not until July 2021 that the VOC gained importance, making it difficult to detect those early clusters with a modest sequencing strategy.

The distribution of the normalised density of clusters is shown in **figure 4** for the whole dataset. Upon downsampling to 50k sequences, we observed a similar pattern of peaks and valleys than with the complete dataset, with some time windows now being *empty* of clusters (e.g., beginning of July 2021, May 2022) (**figure 4**).

**Figure 4.**
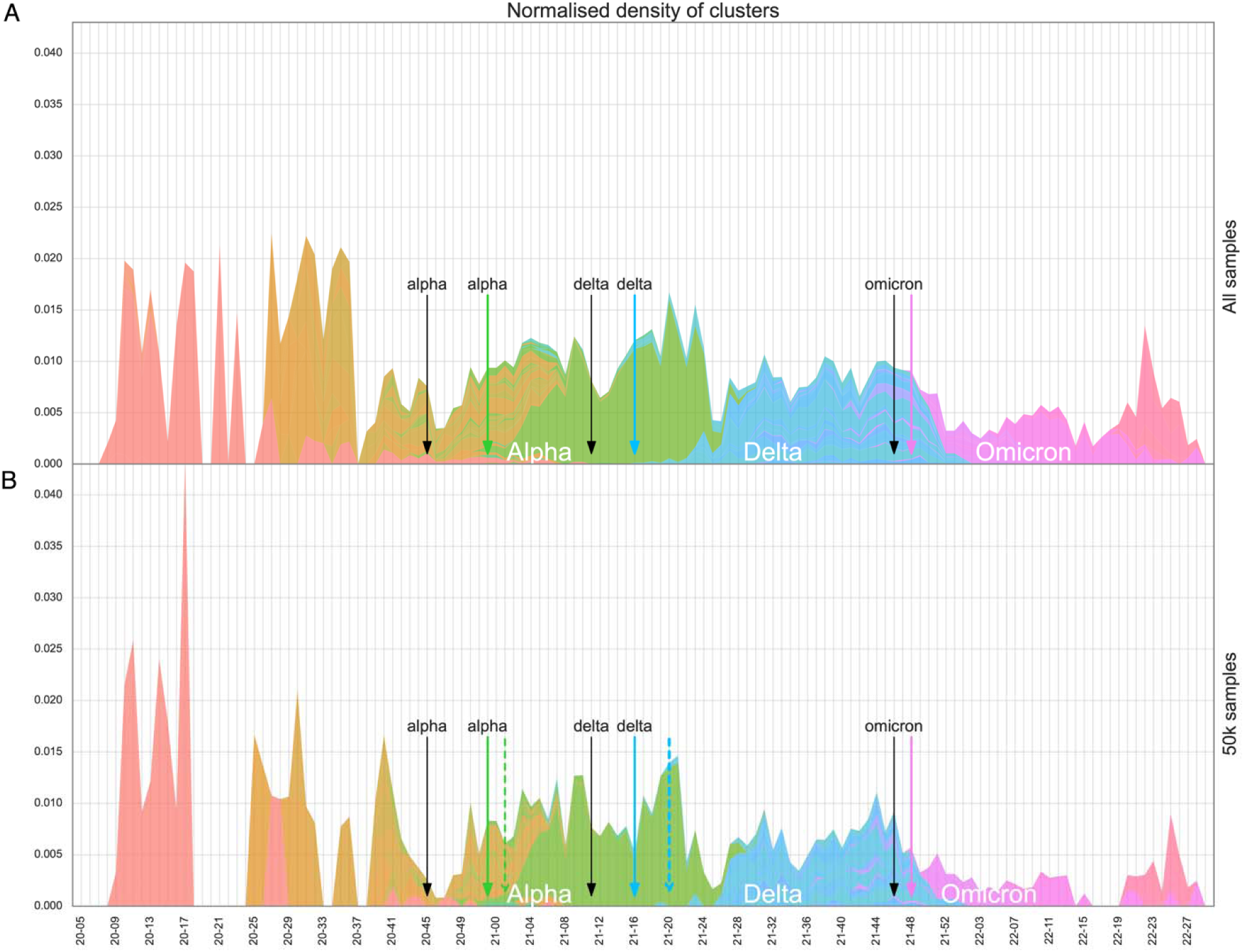
(A) Normalised density of clusters per day for all available sequences. (B) Normalised density of clusters per day for a sample of 50k sequences. The absolute number of active clusters detected each day has been normalised by the number of sequences obtained in a time window of +/− 15 days. The colours indicate the VOC (Alpha in green, Delta in blue, and Omicron in pink). Black arrows indicate the date at which the first sequence of each variant was isolated; coloured arrows highlight the date at which the first cluster of each VOC was detected when considering all 143k samples; coloured dashed arrows show the date at which the first cluster of each VOC would be detected if considering only 50k samples.

### Effect of downsampling on VOC geographic spread and cluster duration

Cluster characteristics such as the cluster duration and geographic spread help public health organisations understand the spread of an outbreak, e.g., a superspreading event, and thereby argue on the introduction of countermeasures. Thus, it is important to study how these cluster characteristics change when reducing the sequencing effort. For all the available genomes, a total of 3,014 clusters were found from which 54 were long distance virus movements (LDVM) (1.8%), defined as events which spread a distance >200 km. The median cluster size included 3 cases (IQR = 2), while for LDVM events included in median 5 cases (IQR = 4.75). On the other hand, for a downsampling size of 50k sequences, 738 clusters were found, from which 17 were LDVM (2.3%). The median number of cases per cluster was 3 cases (IQR = 1), while it was 4 cases (IQR = 2) for LDVM events.

Downsampling had little effect on the duration of clusters, a result consistent for all VOCs. The median duration ranged from 10 days for the complete dataset to 14 days for 2,500 sequences (**figure S8**).

Downsampling resulted in a reduction of the maximum distance within cases of a cluster (**figure S9**). Most clusters were found to be localised in a single canton. Clusters spreading up to five cantons were less frequent, and clusters appearing in six or more cantons were sporadic. A small, yet significant correlation was found between the number of samples in a cluster and the number of affected cantons (**figure S10**). While the absolute number of captured LDVM decreased upon downsampling, their proportion increased, meaning that LDVM got enriched upon downsampling (**figure 5**).

**Figure 5.**
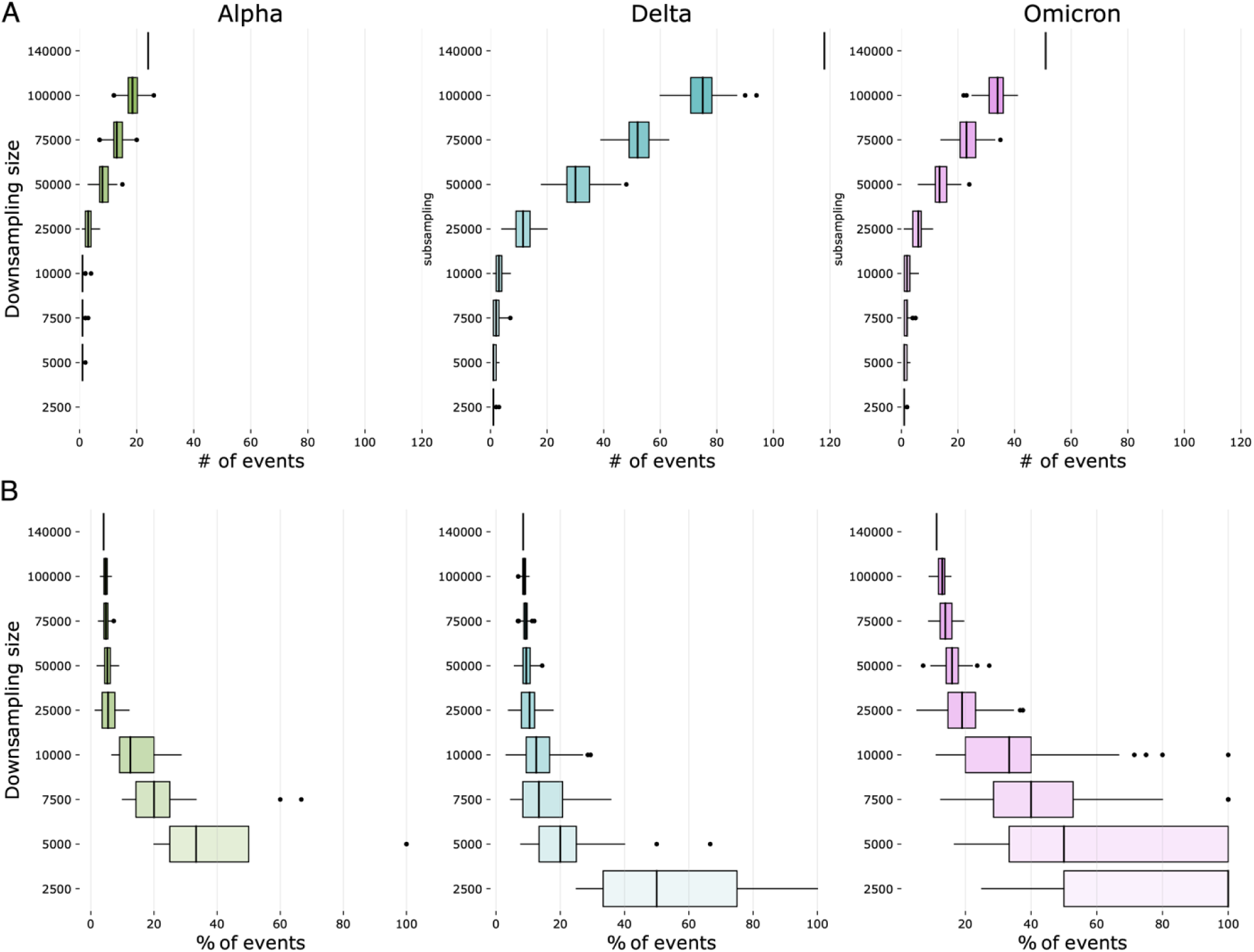
Effect of downsampling on long distance virus movements (LDVM). (A) Absolute number of LDVM detected at each downsampling size. (B) Percentage of clusters that were found to be LDVM at each downsampling size.

## Discussion

Within a surveillance program, sampling strategy and sample selection are important factors for pathogen sequencing as they influence the ability to detect emerging lineages [14] as well as epidemiological parameters and phylodynamic inferences [16–18]. For surveillance of SARS-CoV-2, the European Centre for Disease Prevention and Control (ECDC) recommends a representative sampling approach across geographic locations and demographics for general surveillance for both situation awareness and rare or novel variant detection. This was the chosen approach for Switzerland’s surveillance program. The recommended minimum prevalence to be aimed for in lineage detection is 2.5% [19]. In Switzerland’s case, the target of the surveillance program for much of the period observed in this study was 2,000 sequences/week, which was just above ECDC recommendation (1,522 sequences/week) for very rare lineages (1% prevalence) in periods of high case load (>100,000 cases/week) [19]. Another study estimated that 5% of cases needed to be sequenced to detect emerging lineages at 0.1 to 1% prevalence [20]. This threshold was met for most of the period before and during the official national surveillance program until the beginning of 2022 (on average 9.7%, **figure 1B**). This means Switzerland forms part of only 6.8% of countries that have sequenced at least 5% of cases in the first two years of the pandemic. 45% of countries have sequenced less than 0.5% of cases [14], highlighting the privileged situation for sequencing-based surveillance in Switzerland.

We were able to conduct this study due to the coordinated SARS-CoV-2 surveillance program and the centralised data collection approach via the Swiss Pathogen Surveillance Platform. Such a platform served to coordinate the data sharing following the FAIR principles (findable, accessible, interoperable, and re-usable) [12,13]. A centralised infrastructure for data collection, processing of genome and epidemiological data is of crucial importance during such a public health crisis.

Concurring with this, we find that simulated sequencing effort by downsampling to around a third of the actual extent had overall only a marginal effect on the first detection date of VOCs as well as their clusters, although this was slightly different for certain lineages based on their unique characteristics and the epidemiological backgrounds within which they emerged (in particular Delta). Interestingly, this also holds true for cluster duration (i.e. active transmission chains of highly identical strains). However, estimates of the introduction speed and growth of a VOC, especially for Omicron, as well as the detection of long distance viral movements were more sensitive and reduced sequencing came at the cost of sensitivity.

In this study, we took a retrospective look at all available data, rather than simulating real-time surveillance retrospectively. This pandemic was the first event of this scale where genomic surveillance was applied and required a certain amount of time for political decision making and the subsequent surveillance setup. This period would confound our analysis as we wanted to see by how much sequencing effort could have been reduced without affecting certain surveillance outcomes.

For this reason, a critical factor for timely lineage detection not assessed in our study is turnaround time (TAT, i.e. the time lag between sample collection and submission of the genome sequence to a surveillance database such as SPSP or GISAID) [21]. The median TAT in Switzerland was 18 days (with IQR of 14 days) over the period of the official national surveillance program. Assuming a TAT of 21 days, the probability of lineage detection before it reaches 100 cases was estimated as 0.51 and 0.96 if 1% or 5% of cases were sequenced, respectively, as simulated with data from Denmark [14]. This highlights that other important factors than just the amount of sequencing should also be targets of surveillance optimisation.

We showed that the effect of sequencing effort differs for different surveillance outcomes. A national surveillance program needs to strike a balance between societal benefits and program costs. To achieve that, desired outcomes of surveillance should be clearly defined and targets set accordingly.

## Acknowledgment

We want to thank the following scientists and technicians at the institution.

University Hospital Basel: Dr. Helena Seth-Smith, Dr. Madlen Stange, Dr. Alfredo Mari, Elisabeth Schultheis, Daniel Gander, Magdalena Schneider, Rosamaria Vesco, and Valerie Courtet for contributing to the SARS-CoV-2 sequencing program.

Institute of Medical Virology, University of Zurich: Stefan Schmutz, Gabriela Ziltener, Nadine Rist, Andrea Hafner, Dr. Maryam Zaheri and Dr. Kevin Steiner for contributing to the SARS-CoV-2 sequencing program.

All individuals involved in the ETH-led sequencing effort https://bsse.ethz.ch/cevo/research/sars-cov-2/swiss-sars-cov-2-sequencing-consortium.html.

## Funding

The Swiss SARS-CoV-2 sequencing program was financed by the Federal Office of Public Health. The SPSP was financed by multiple grants: NRP72 program (407240_177504; AE), swissuniversities openscience (AE/AN), Swiss National Science Foundation (310030_192515; AE), Bangerter Rhyner Foundation (AE/AN), SERI (AN), Federal Office of Public Health (AE/AN).

## Ethical statement

The study was conducted as part of the routine SARS-CoV-2 genomic surveillance program. SPSP has ethical approval with the IRB number 2019-01291.

## Conflict of interest

None.

## Authors’ contributions

FW and BCG analysed the data. BCG developed the cluster detection tool. AE and AN designed and supervised the study. FW, BCG, AE and AN contributed to writing the manuscript. All other authors contributed data as part of the Swiss surveillance program and reviewed the manuscript. Except for co-first and co-last authors, names are sorted alphabetically.

## Data availability

Data, scripts and software are available under

https://gitlab.uzh.ch/appliedmicrobiologyresearch/amr_publications/spsp_paper

https://gitlab.sib.swiss/SPSP/sarscov2/cluster-detection/-/tree/master/paper

https://gitlab.sib.swiss/SPSP/sarscov2/cluster-detection

## Methods

### Data

All sequences shared with SPSP (https://www.spsp.ch) until the 5^th^ of August 2022 were used (n=143,260) in this study. Genomes sequenced with the explicit purpose of outbreak investigation were removed. The earliest genome collection date was 24 February 2020, the latest 1 August 2022. Genomes with incorrect collection dates (obvious data entry errors) were excluded. Pango lineage assignments were performed on SPSP in regular intervals and correspond in the dataset to the latest Pangolin version available on 5 August 2022 [22]. Additionally, Swiss COVID-19 case data were taken from the FOPH dashboard [https://www.covid19.admin.ch/, last accessed 20 September 2022].

### Downsampling and Key Measures

The complete dataset was downsampled to sizes ranging from 2,500 to 100,000 using random uniform sampling and 100 iterations. VOC labels (Alpha, Delta, and Omicron) were given to all genomes with assigned Pango sublineages associated with a given VOC. The introduction date was determined as the earliest collection date of a given VOC in the dataset. The speed of introduction was calculated as the slope of a linear regression model of the log of exponential growth phase of a given VOC. The end of the exponential growth phase was determined based on the complete dataset as 2021-03-01 for Alpha, 2021-08-01 for Delta, and 2022-01-01 for Omicron.

The lineage diversity was calculated as the Shannon diversity index [23] using the R package vegan [24]. It accounts for both lineage richness and evenness, i.e., the number of lineages as well as their proportion. For example, the diversity in two samples with the same number of lineages would be greater in the sample with even proportions for each lineage compared to the sample with a few dominant and many rare lineages.

The nucleotide diversity was calculated for whole genome alignments (produced with mafft v7.505 [26]) of all sequences per calendar week using the R package pegas [25].

### Cluster Definition

A three-step method was used to identify sequence clusters. First, sequences with 0 single nucleotide polymorphism (SNP) distance (including missing regions) were grouped to obtain mutually-exclusive clusters. These early clusters should have a minimum of 3 sequences. At this point, if a SNP difference occurred in a region that was missing in an otherwise identical sequence, this was counted as a SNP difference of 1, and the two sequences would be placed in different clusters. To overcome this and account for the possible mutations within a transmission chain, 0-SNP-clusters were expanded by adding sequences at 1 SNP distance. This resulted in multiple non-mutually-exclusive clusters with overlapping sequences. Finally, the expanded clusters sharing sequences were merged using hierarchical clustering to create mutually-exclusive super-clusters. To ensure the quality of the super-clusters, super-clusters containing sequences isolated more than 30 days apart were removed from the study. This time window was selected based on the estimated mutation rate of SARS-CoV-2 of 1.1LJ×LJ10^−3^ subs/site/year (i.e., around two to three mutations per month) [27]. To maximise the number of super-clusters, the duration of 0-SNP-clusters was studied by comparing the number of sequences, cluster counts, and percentage of bad clusters for different duration thresholds (**figure S11**). A threshold of 20 days for the 0-SNP-clusters was found to maximise the number of sequences analysed, the number of clusters found and to minimise the resulting number of erroneous clusters.

### VOC cluster detection dates and geographic spread

The delay in VOC cluster detection was computed in relation to the first detection of a VOC sequence. The effect of downsampling on the detection of the first VOC clusters was analysed by finding the distribution of the first cluster detection dates in the downsampled datasets. The downsample size of 50k samples was considered the smallest size in which the VOC detection dates could be retrieved without major delays. Therefore, this size was selected as the downsample size for further analysis.

Clusters containing the same sequences in the complete dataset and a downsampled dataset (50k sequences) were matched. Discrepancies in cluster matching can arise because 1) large clusters lasting longer than our 30-day-threshold are discarded but are found in part in the downsampled dataset, or 2) larger clusters from the complete dataset were split into several clusters of shorter duration in the downsampled dataset given that the common sequences are not in the sample anymore.

The distance between the cantons affected by clusters was computed as the Euclidean distance in kilometres between the capital of each canton.

### Normalised density of clusters per day

Clusters were determined for the complete dataset as well as the 50k downsampled dataset (see above). A cluster was considered active during the period between the isolation of its first and last sequences. Given that the sequencing effort was not consistent during the study period, only considering the absolute counts of clusters per day would introduce a bias towards the periods with more sequencing effort. Therefore, the absolute number of clusters active each day was normalised by the number of sequences isolated in a time window of ± 15 days (30 days being the estimated maximum duration of a cluster).

## Supplemental Information

**Figure S1.**
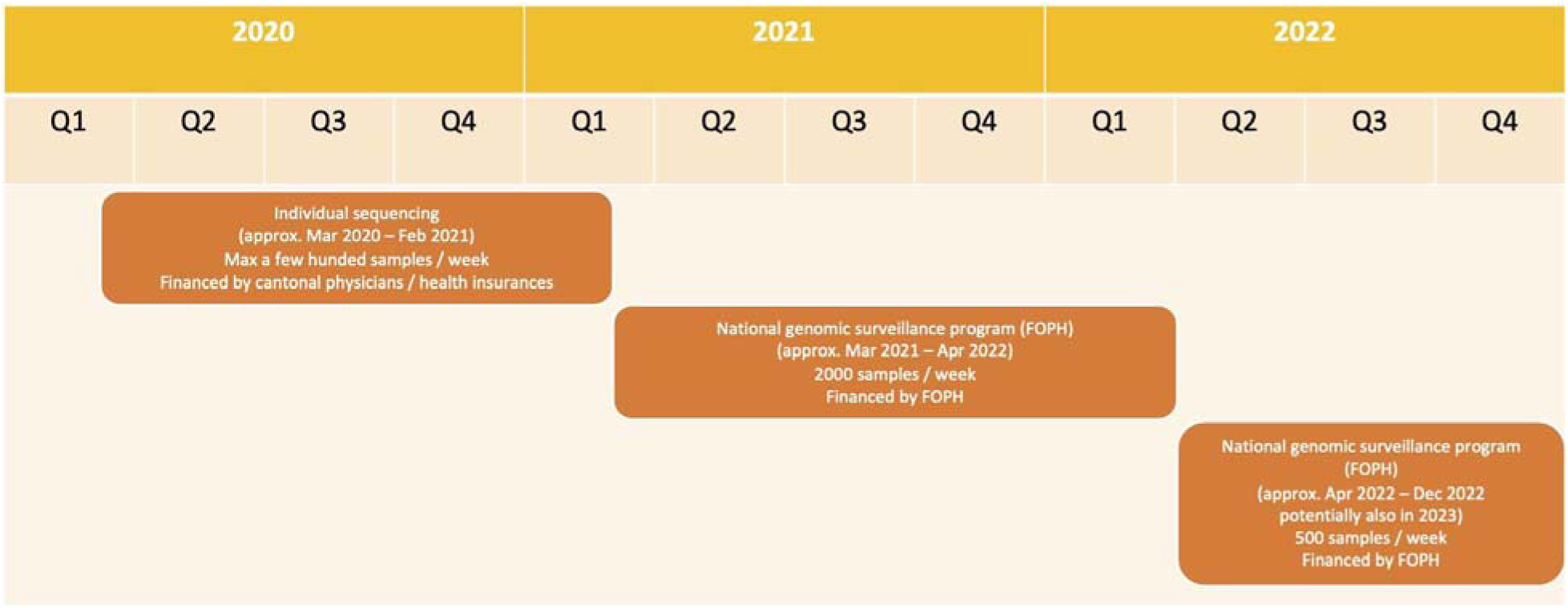
Timeline of the Swiss SARS-CoV-2 surveillance sequencing.

**Figure S2.**
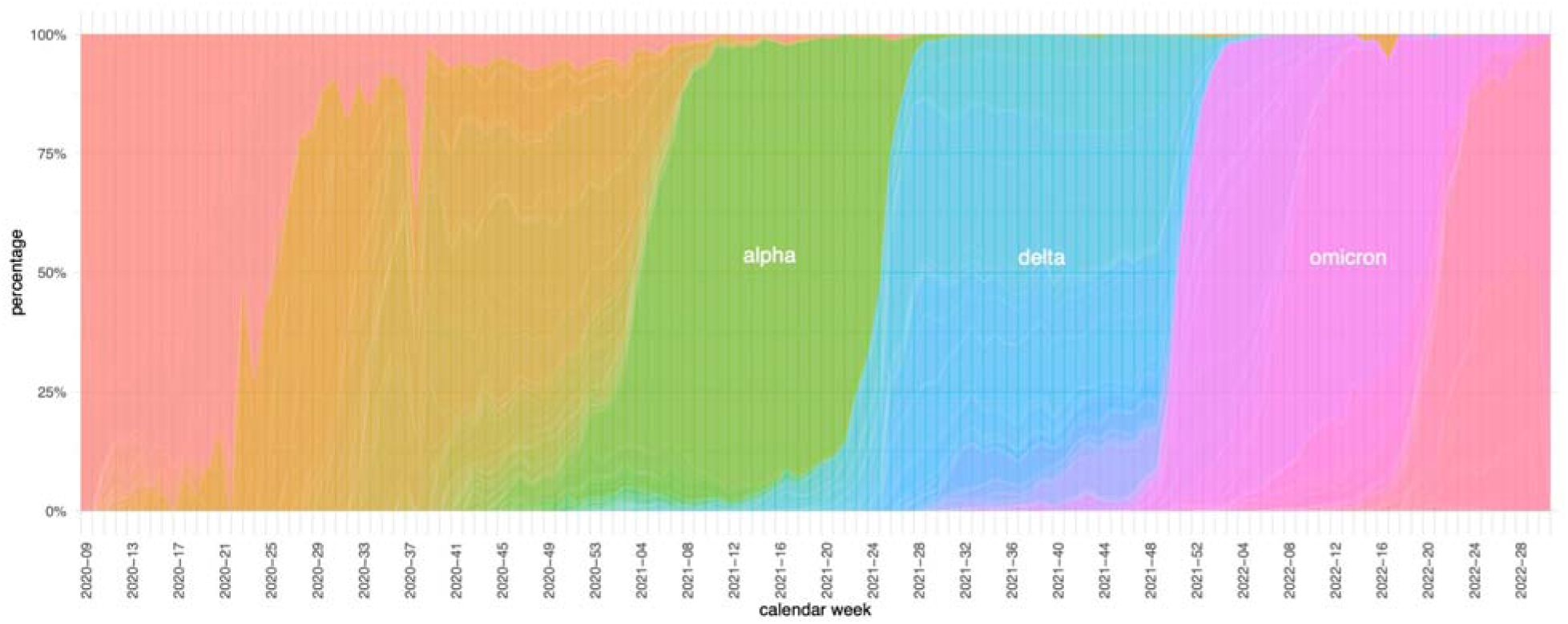
Proportional plot of the sequenced lineages (as shown in figure 1C).

**Figure S3.**
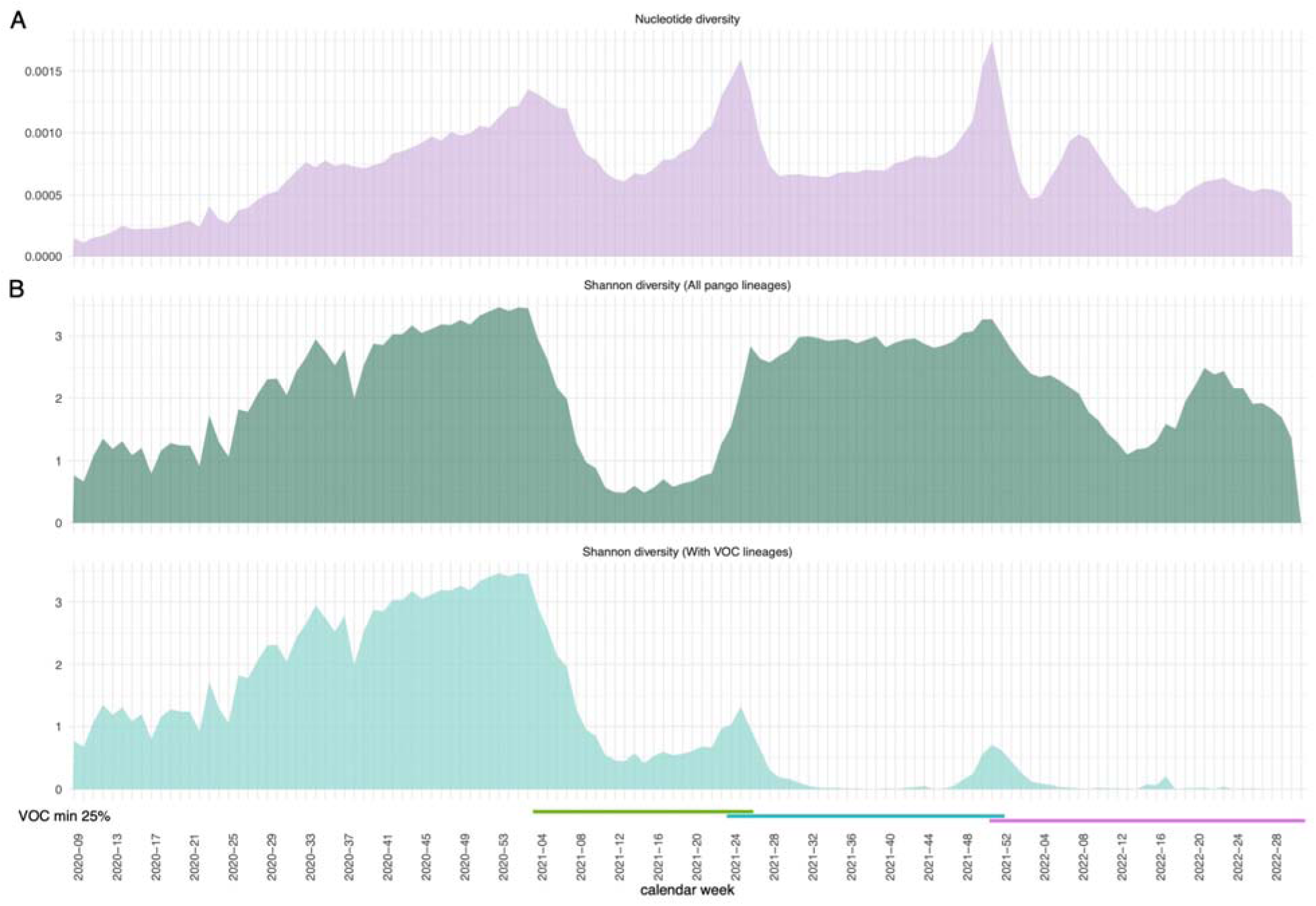
(A) The genetic diversity per calendar week as measured by nucleotide diversity and (B) lineage diversity per calendar week expressed by Shannon diversity. The diversity was calculated using either counts for all Pango lineages (top) or consolidating all sublineages belonging to the VOCs Alpha, Delta, and Omicron into one lineage, respectively (bottom). The periods in which each VOC had at least 25% prevalence is indicated by the coloured bars (green: Alpha; blue: Delta; Pink: Omicron)

**Figure S4.**
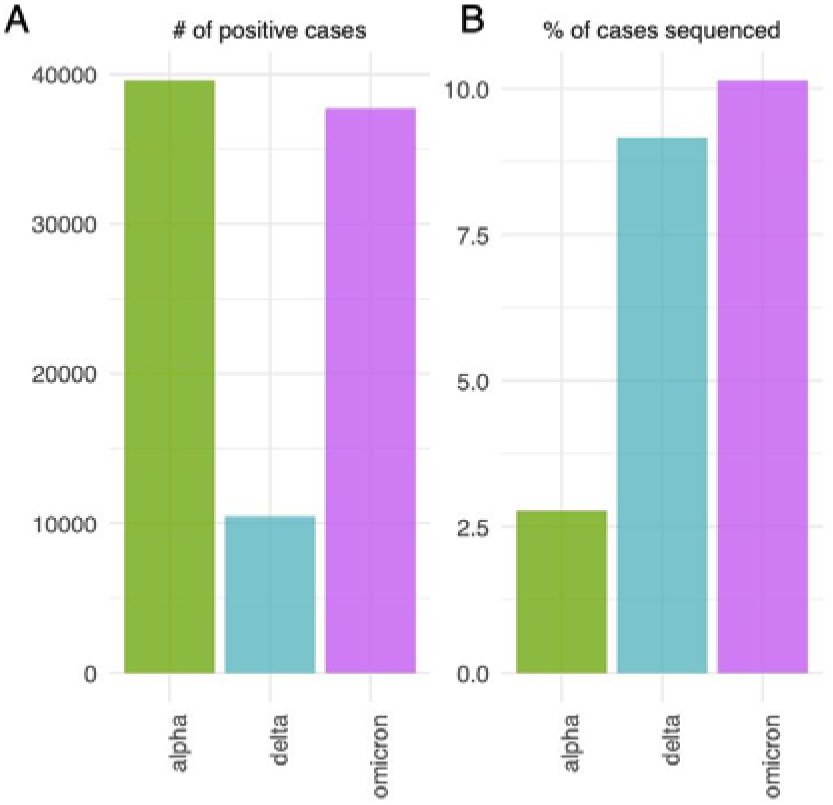
(A) Number of positive cases and (B) percentage of sequenced cases, each at the time (calendar week) of the first detection of a given VOC.

**Figure S5.**
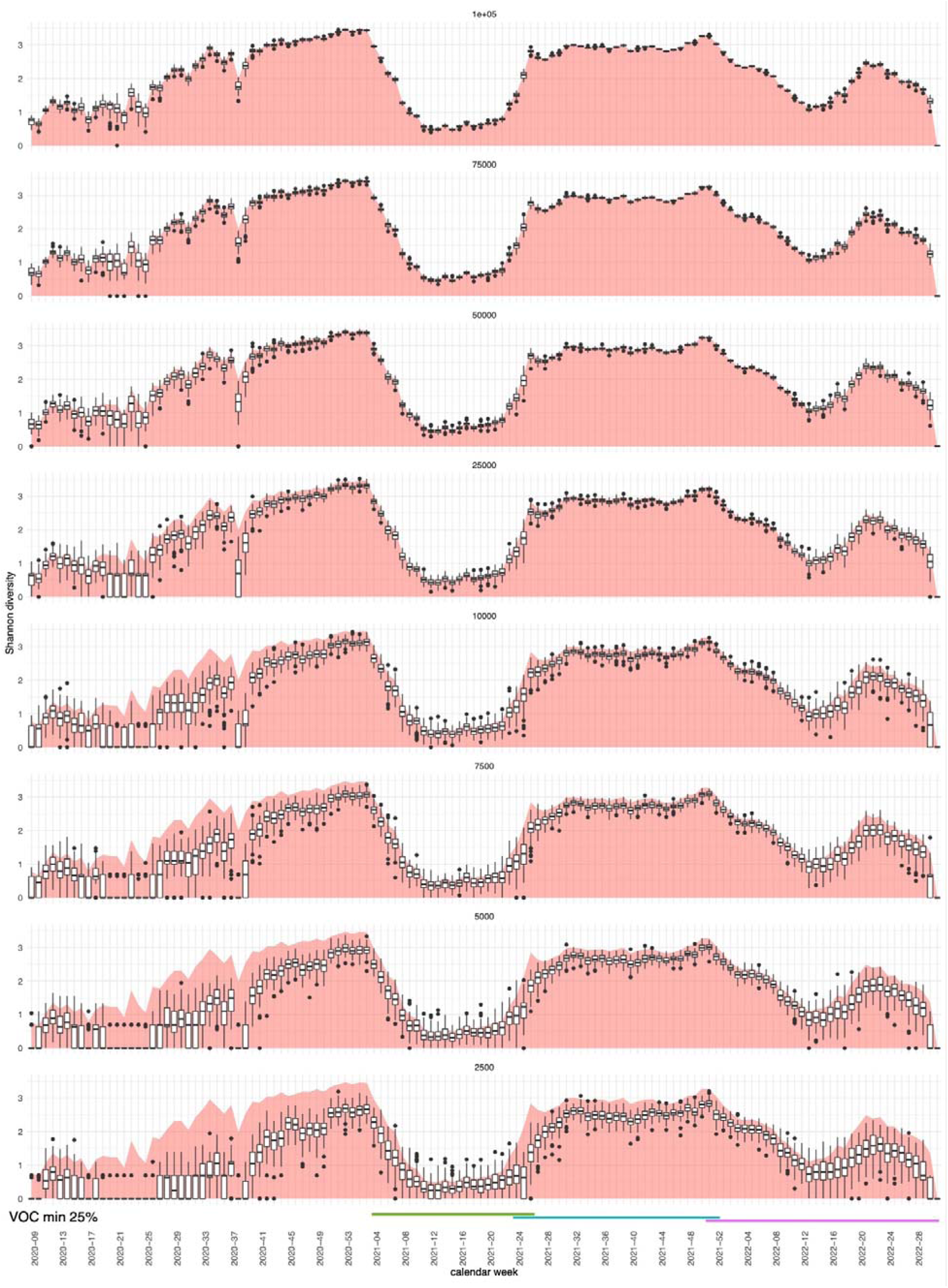
The lineage diversity per calendar week expressed by Shannon diversity upon downsampling (with 100 iterations) using all Pango lineages. The red plot in the background shows the diversity of the complete dataset as a reference (cf. suppl. figure S1). The periods in which each VOC had at least 25% prevalence is indicated by the coloured bars (green: Alpha; blue: Delta; Pink: Omicron)

**Figure S6.**
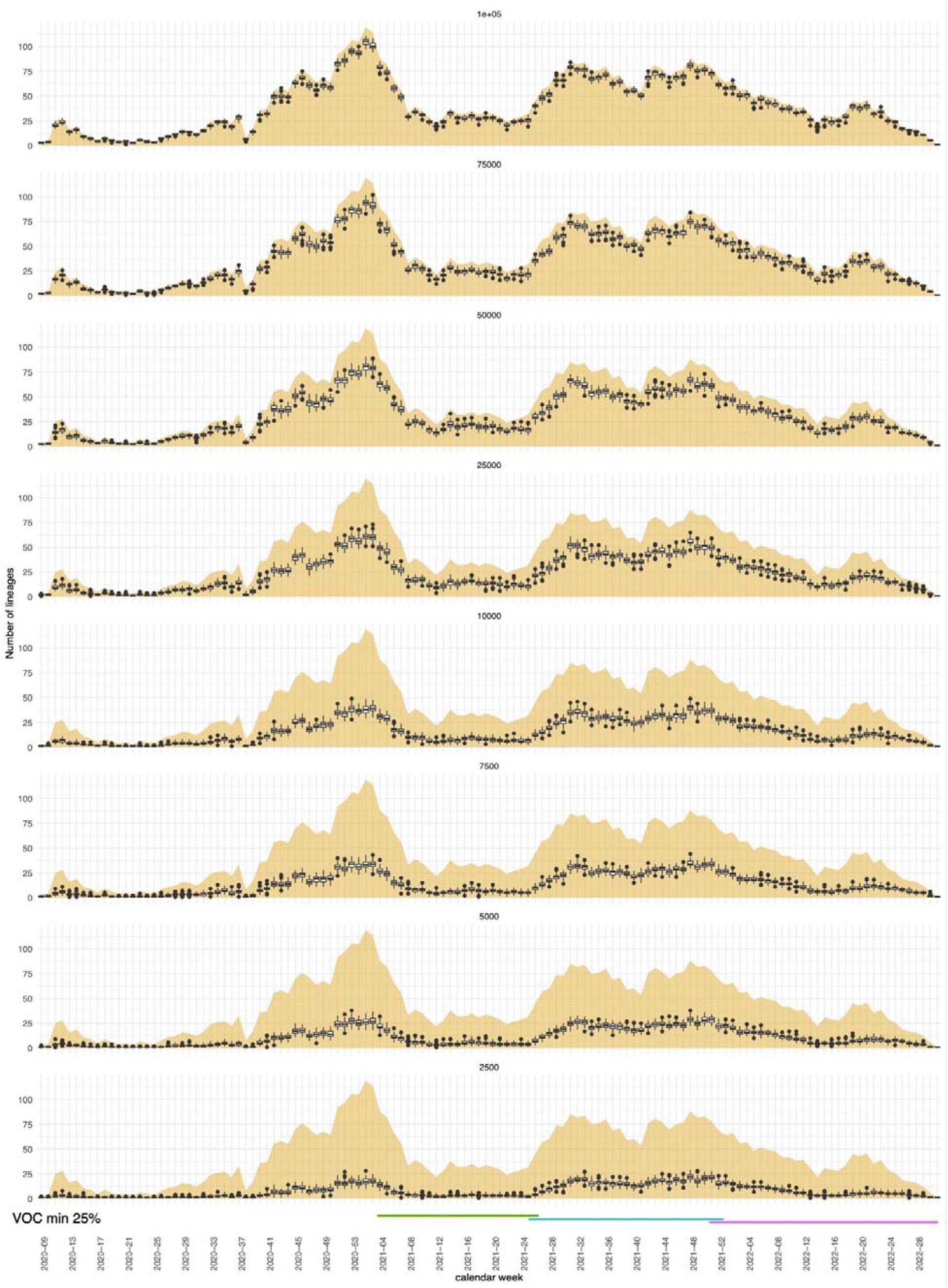
The number of lineages detected per calendar week upon downsampling (with 100 iterations) using all Pango lineages. The yellow plot in the background shows the number of lineages of the complete dataset as a reference. The periods in which each VOC had at least 25% prevalence is indicated by the coloured bars (green: Alpha; blue: Delta; Pink: Omicron)

**Figure S7.**
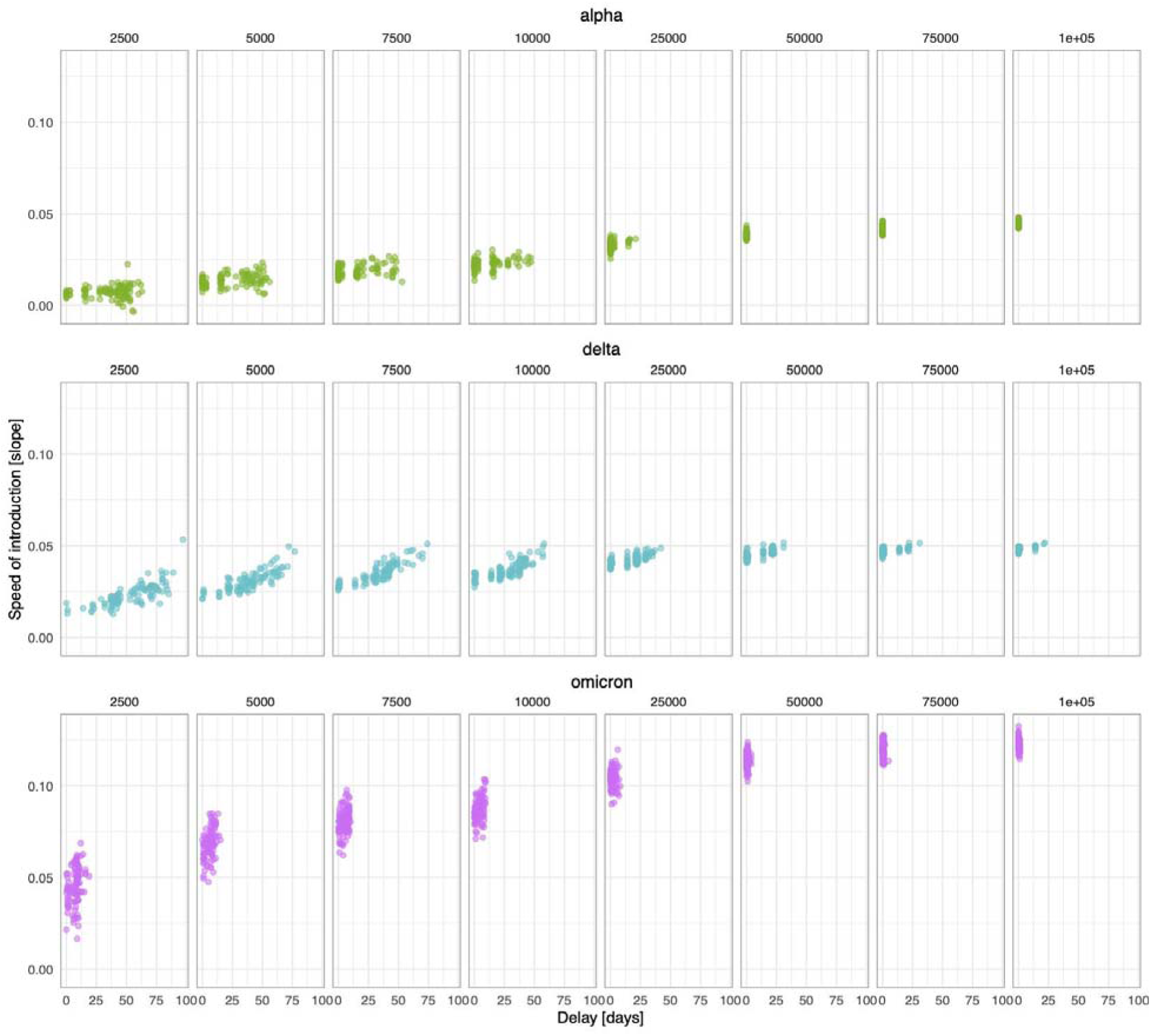
Effect of downsampling on speed of introduction and delay of first detection of VOCs plotted against each other for each VOC.

**Figure S8.**
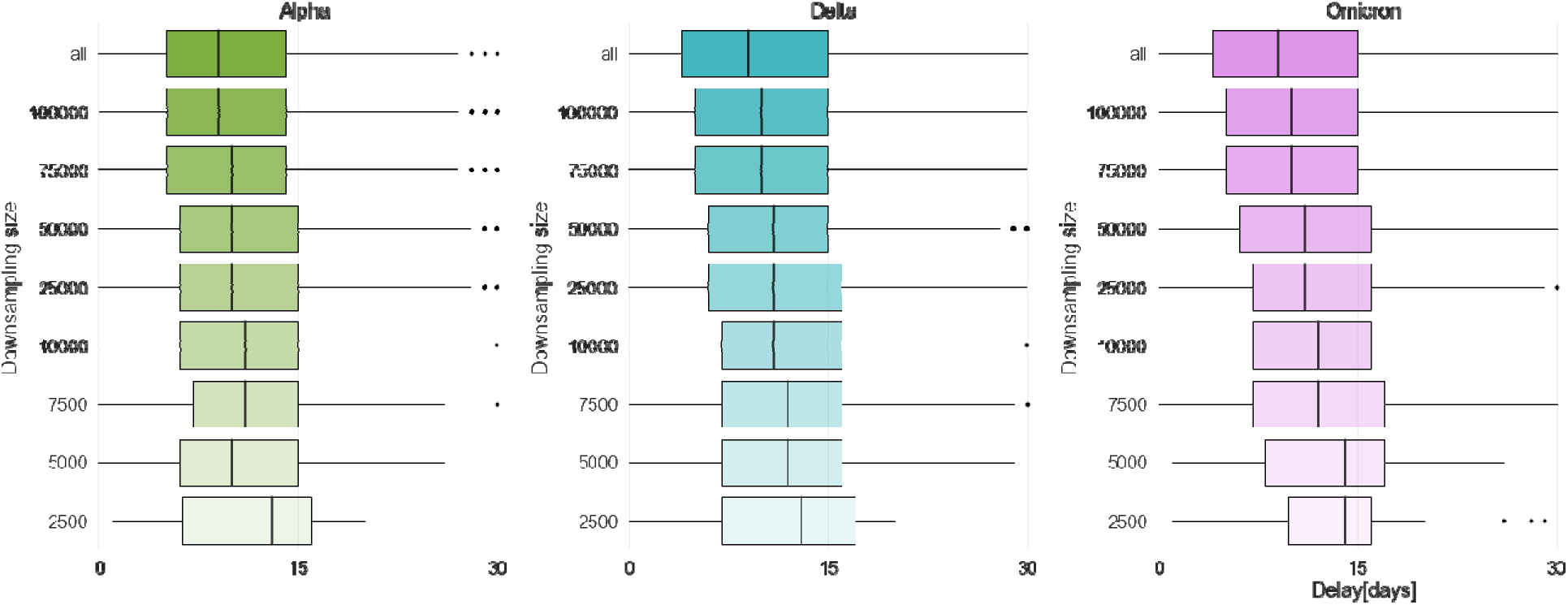
Effect of downsampling on cluster duration. Each boxplot represents the duration in days of the clusters for different downsampling sizes. The delays are expressed as a distribution given that 100 random iterations were taken for each downsampling. Colour code: delays for VOC Alpha in green, for VOC Delta in blue, and for VOC Omicron in pink.

**Figure S9.**
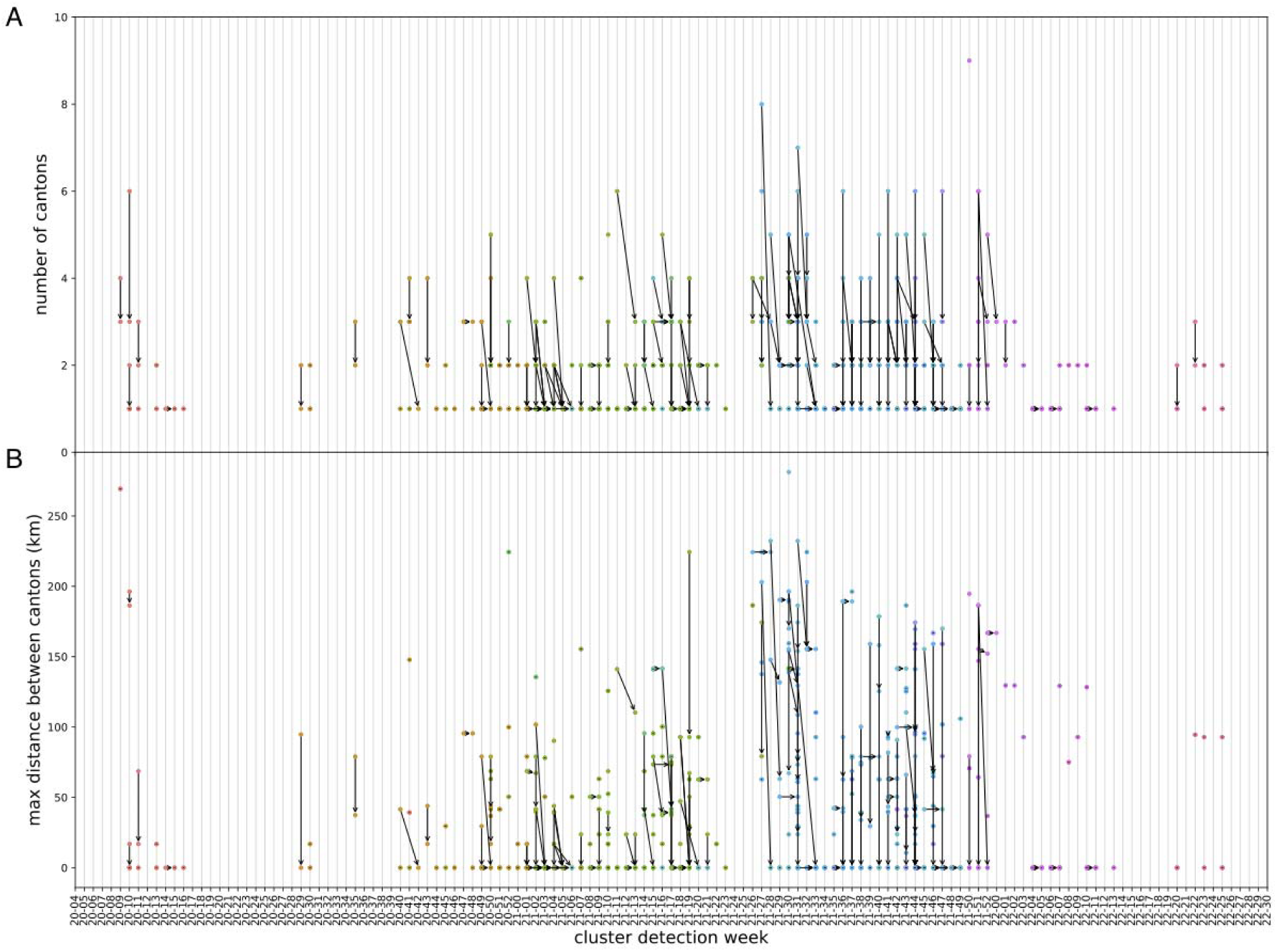
Shift in geographic spread of clusters after downsampling. (A) Geographic spread represented by the number of cantons affected by a cluster. (B) Geographic spread represented by the maximum distance between the cantons affected by each cluster. The dots represent clusters coloured by VOC (Alpha in green, Delta in blue, and Omicron in pink). Dots connected with an arrow represent the same cluster before and after downsampling (143k to 50k samples). Dots without an arrow indicate the clusters for which no shift occurred. Unsuccessfully matched clusters are not represented. The arrow direction indicates the shift in number of affected cantons and the cluster detection date after downsampling.

**Figure S10.**
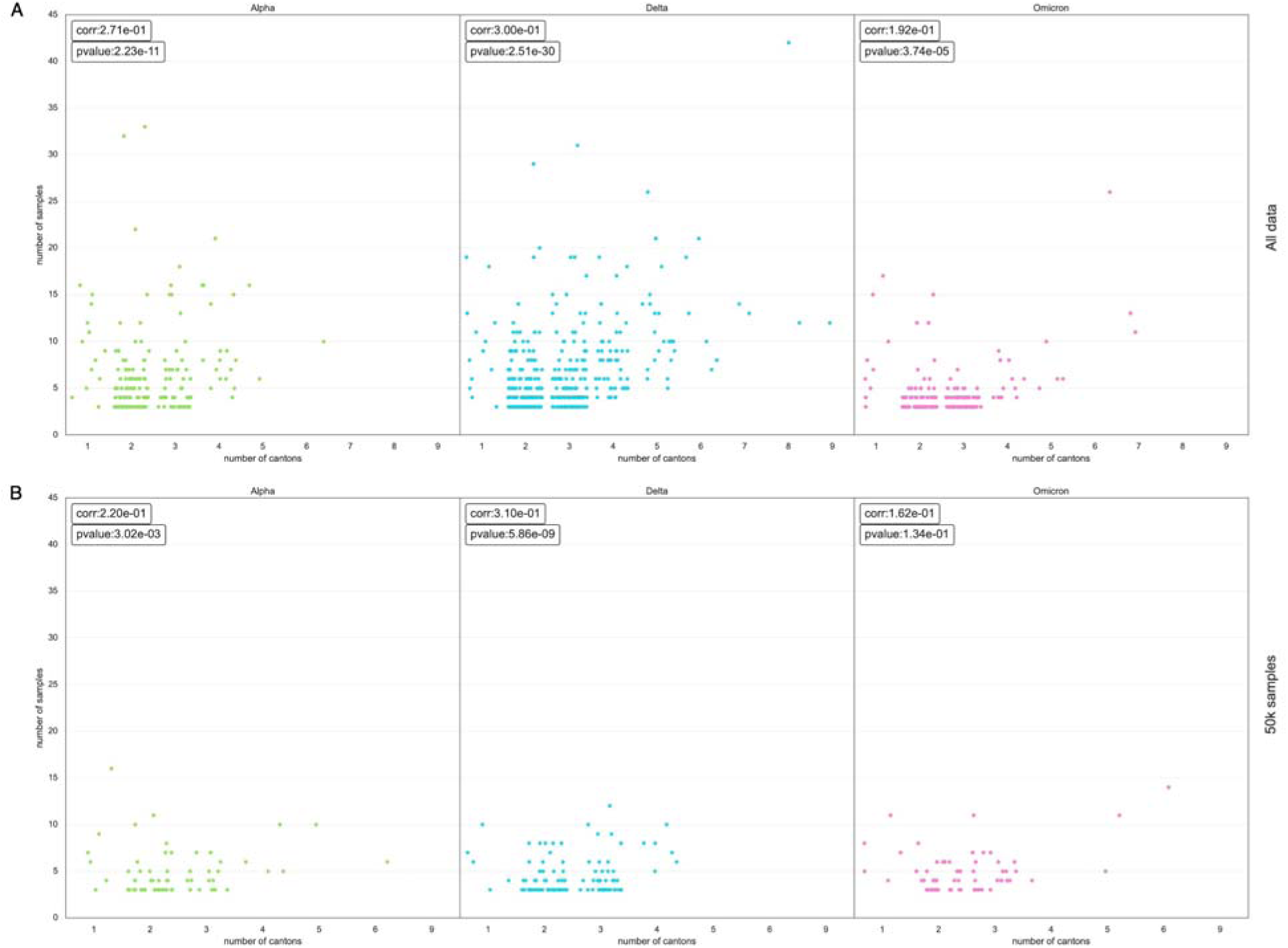
Correlation between cluster size and number of cantons affected per VOC. We show the correlation between the number of samples in each cluster and the number of cantons affected in the cluster. (A) The results were obtained when considering all the samples in the SPSP dataset. (B)The results obtained when using 50k samples. Each panel represents a VOC.

**Figure S11.**
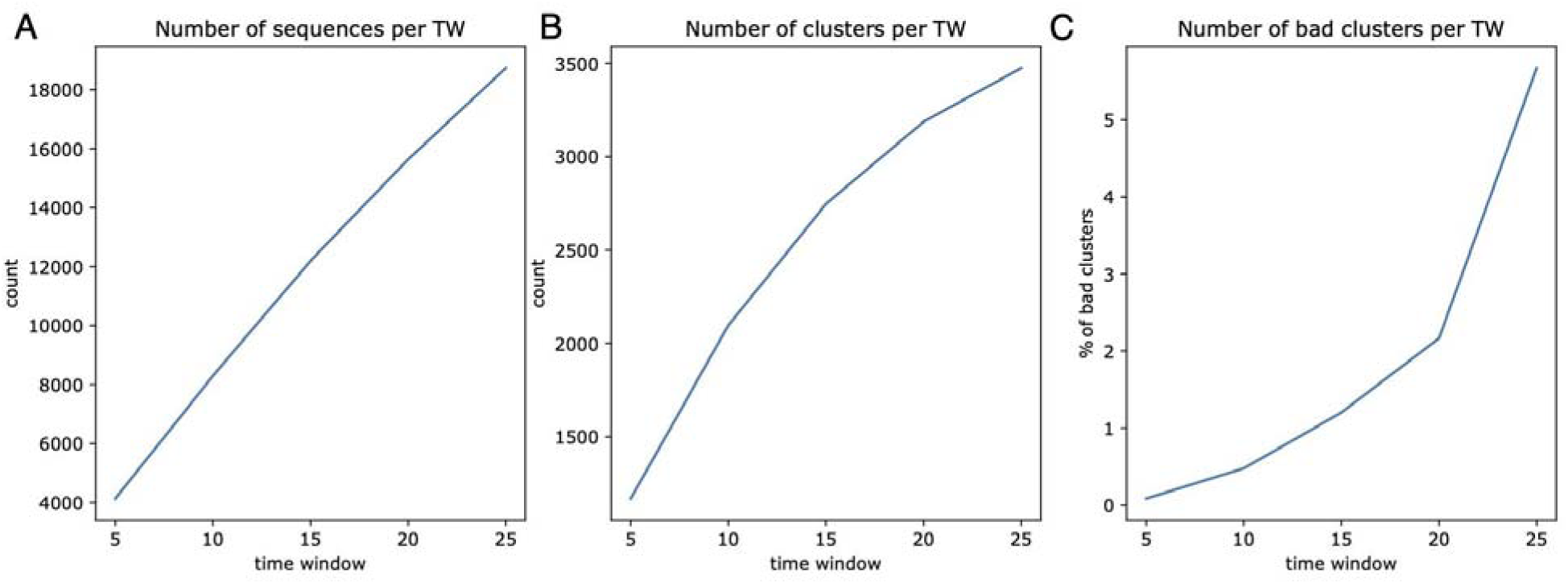
Analysis of 0-SNP cluster time-windows. (A) Number of sequences analysed per time window. (B) Number of superclusters found for each 0-SNP cluster time window. (C) Number of superclusters spanning more than 30 days for each 0-SNP cluster time window.

## Notes

### Competing Interest Statement

The authors have declared no competing interest.

### Author Declarations

Ethics committee/IRB of Ethikkommission Nordwest-und Zentralschweiz (EKNZ) name gave ethical approval for this work

