## Supplemental Information for "How much should we sequence? An analysis of the Swiss SARS-CoV-2 surveillance effort"

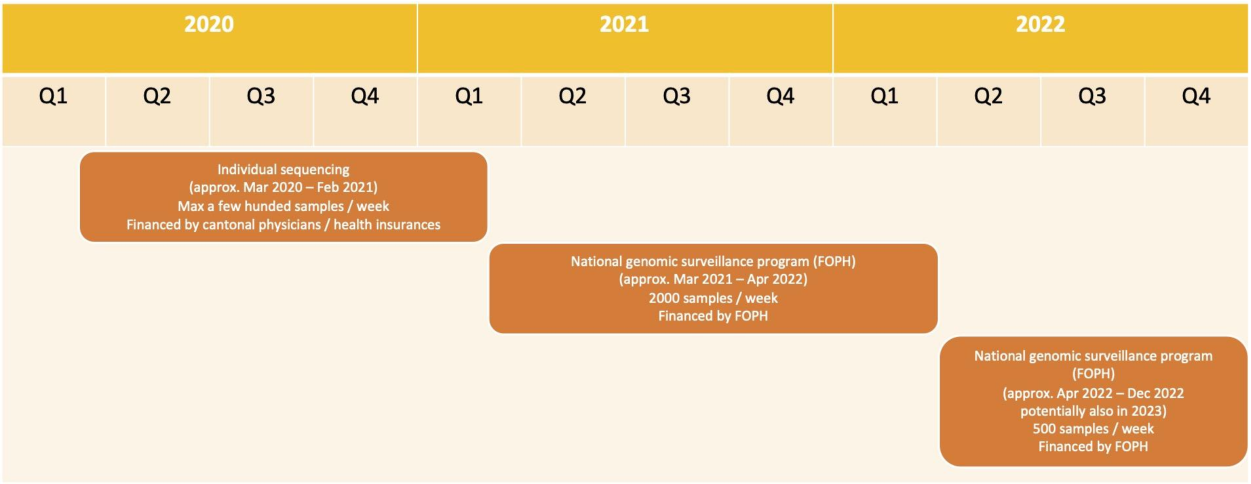


**Figure S1** Timeline of the Swiss SARS-CoV-2 surveillance sequencing.

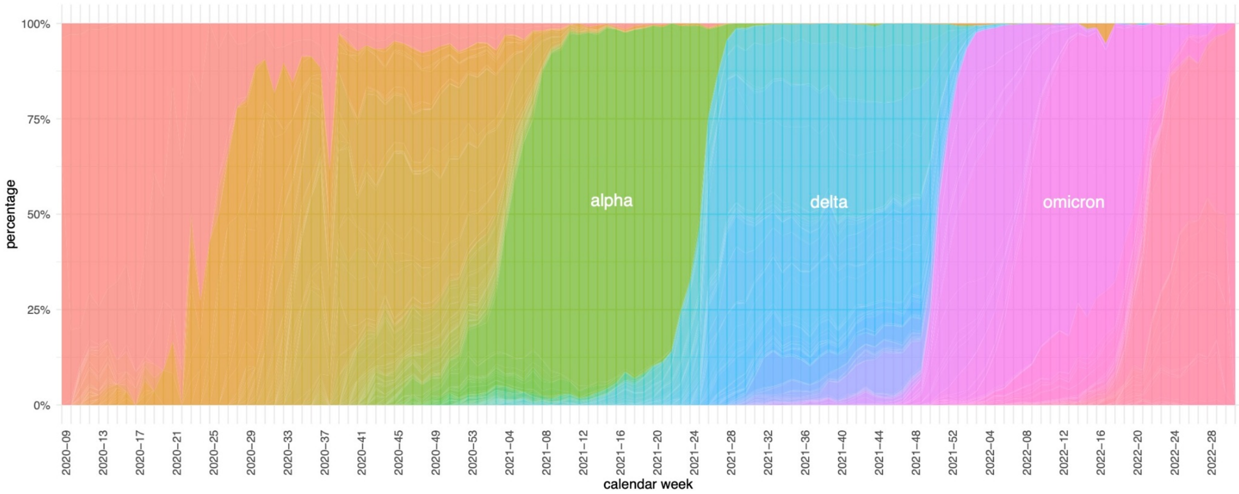


**Figure S2** Proportional plot of the sequenced lineages (as shown in figure 1C).

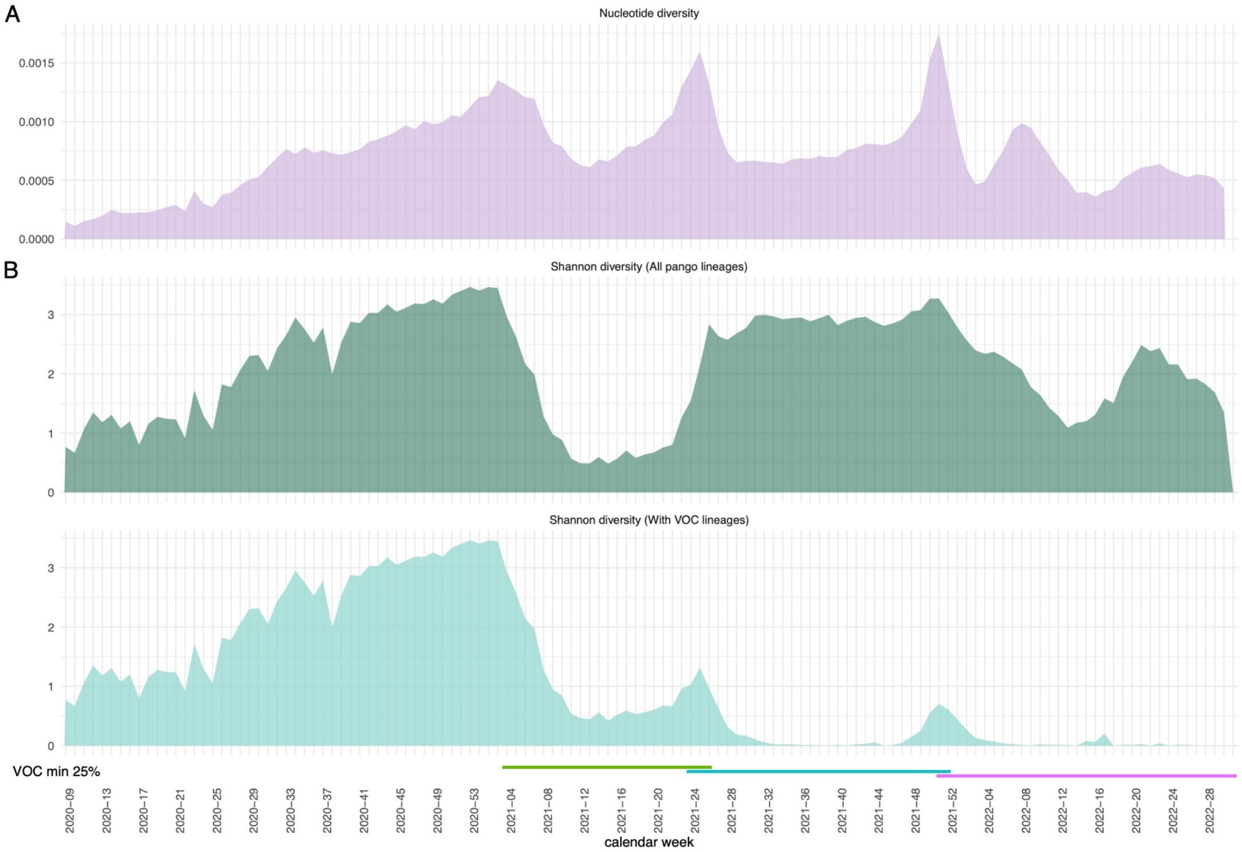


**Figure S3** (A) The genetic diversity per calendar week as measured by nucleotide diversity and (B) lineage diversity per calendar week expressed by Shannon diversity. The diversity was calculated using either counts for all Pango lineages (top) or consolidating all sublineages belonging to the VOCs Alpha, Delta, and Omicron into one lineage, respectively (bottom). The periods in which each VOC had at least 25% prevalence is indicated by the coloured bars (green: Alpha; blue: Delta; Pink: Omicron)

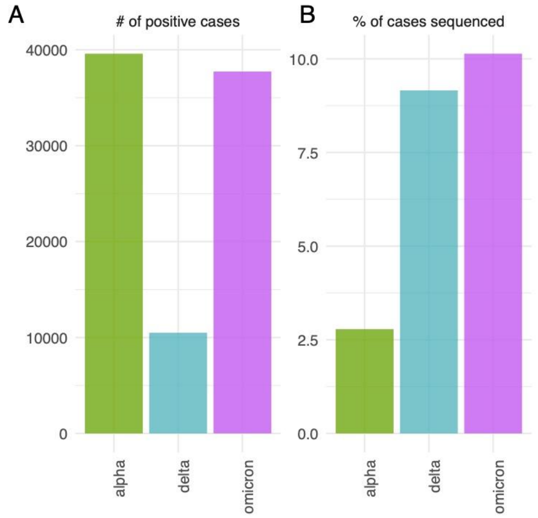


**Figure S4** (A) Number of positive cases and (B) percentage of sequenced cases, each at the time (calendar week) of the first detection of a given VOC.

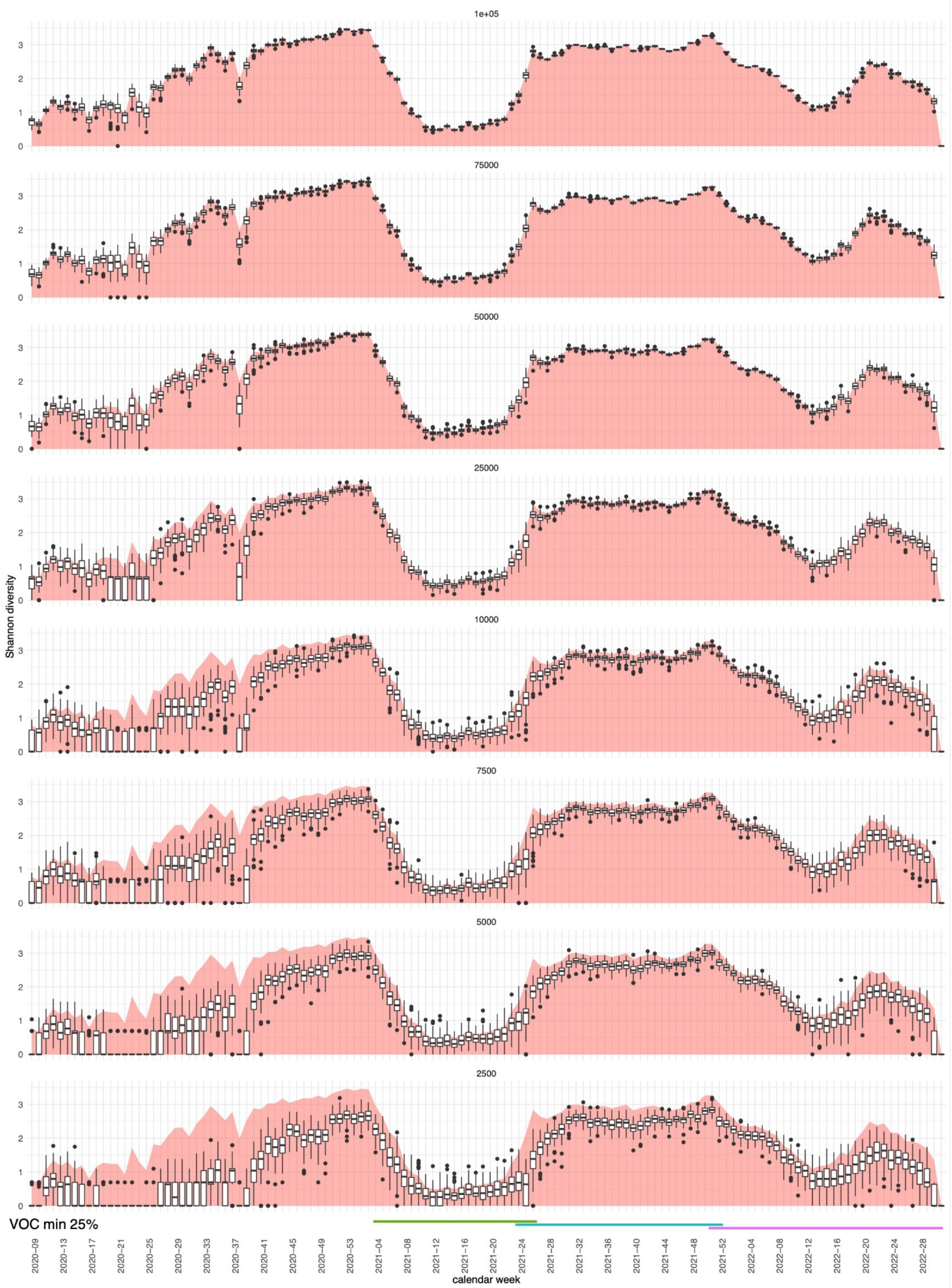


**Figure S5** The lineage diversity per calendar week expressed by Shannon diversity upon downsampling (with 100 iterations) using all Pango lineages. The red plot in the background shows the diversity of the complete dataset as a reference (cf. suppl. figure S1). The periods in which each VOC had at least 25% prevalence is indicated by the coloured bars (green: Alpha; blue: Delta; Pink: Omicron)

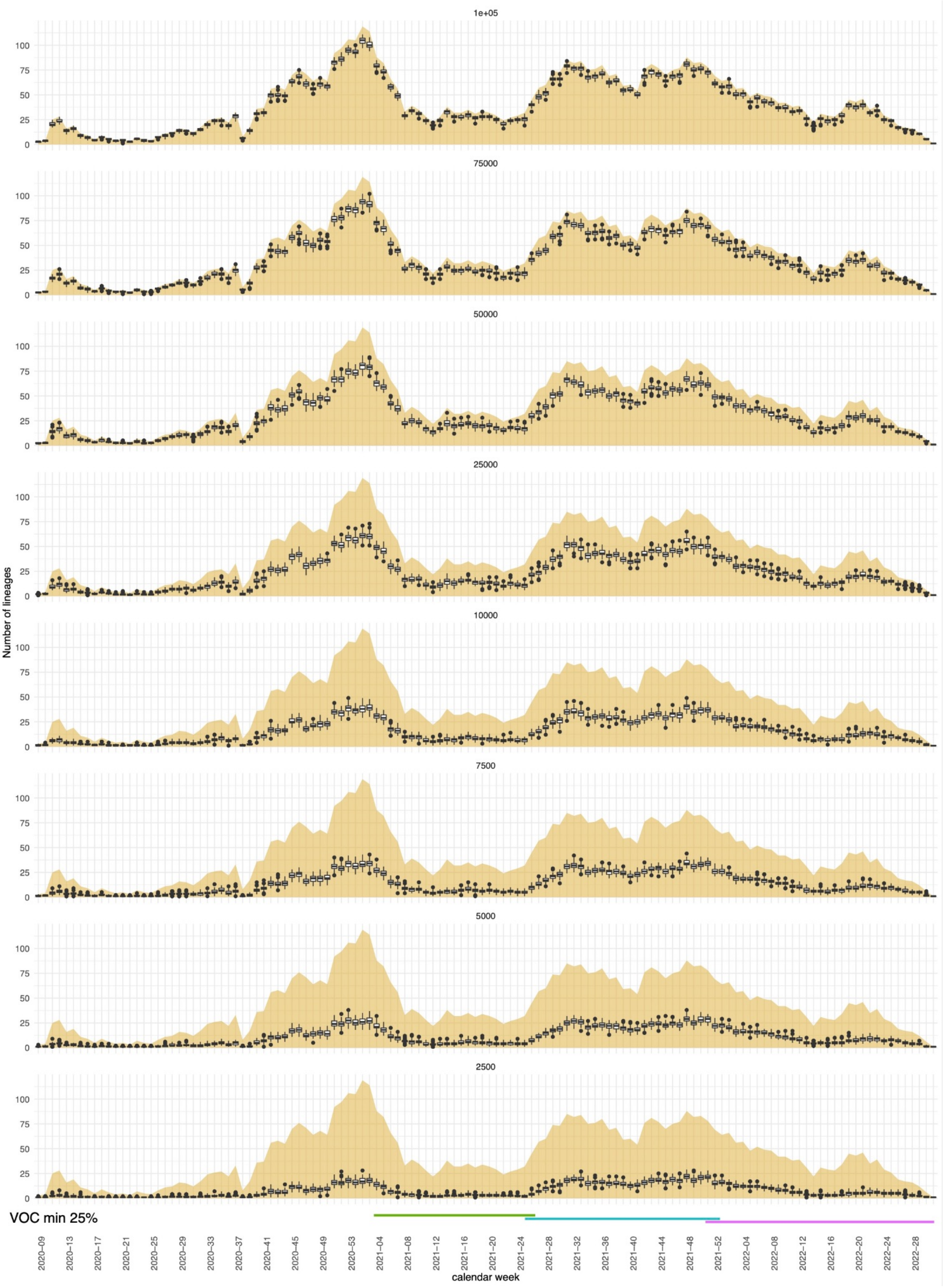

**Figure S6** The number of lineages detected per calendar week upon downsampling (with 100 iterations) using all Pango lineages. The yellow plot in the background shows the number of lineages of the complete dataset as a reference. The periods in which each VOC had at least 25% prevalence is indicated by the coloured bars (green: Alpha; blue: Delta; Pink: Omicron)

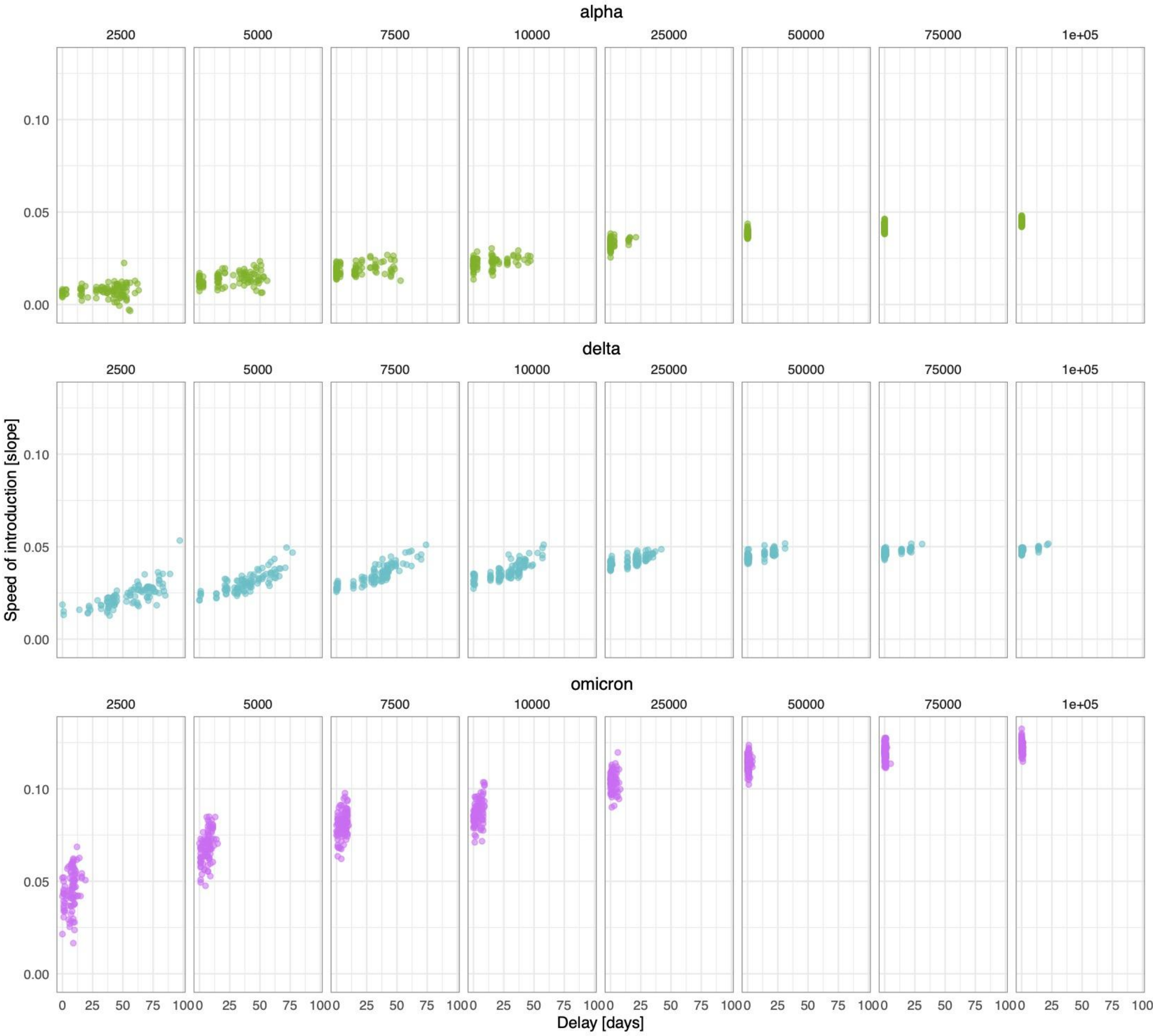


**Figure S7** Effect of downsampling on speed of introduction and delay of first detection of VOCs plotted against each other for each VOC.

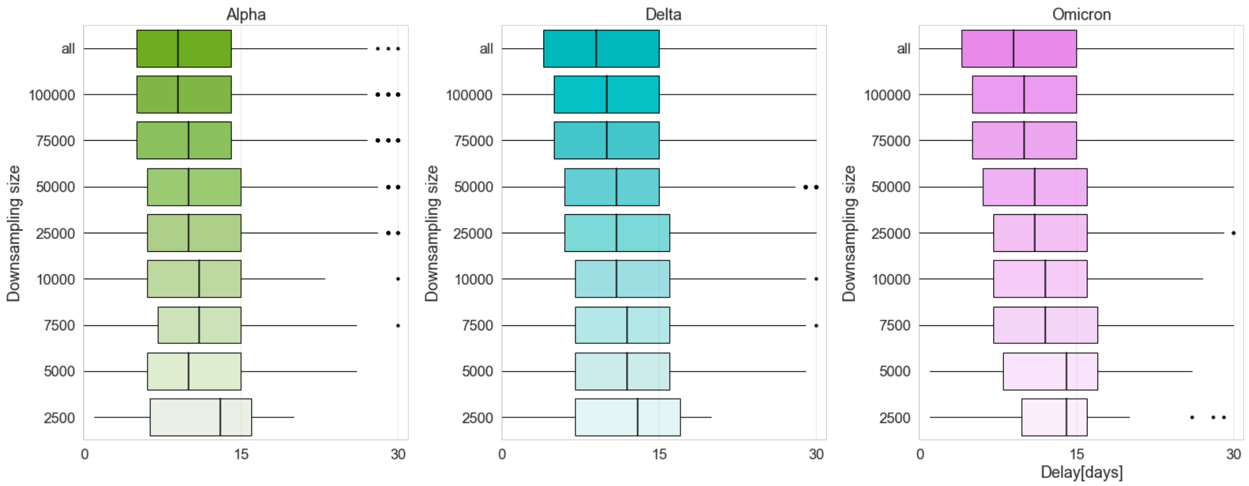


**Figure S8** Effect of downsampling on cluster duration. Each boxplot represents the duration in days of the clusters for different downsampling sizes. The delays are expressed as a distribution given that 100 random iterations were taken for each downsampling. Colour code: delays for VOC Alpha in green, for VOC Delta in blue, and for VOC Omicron in pink.

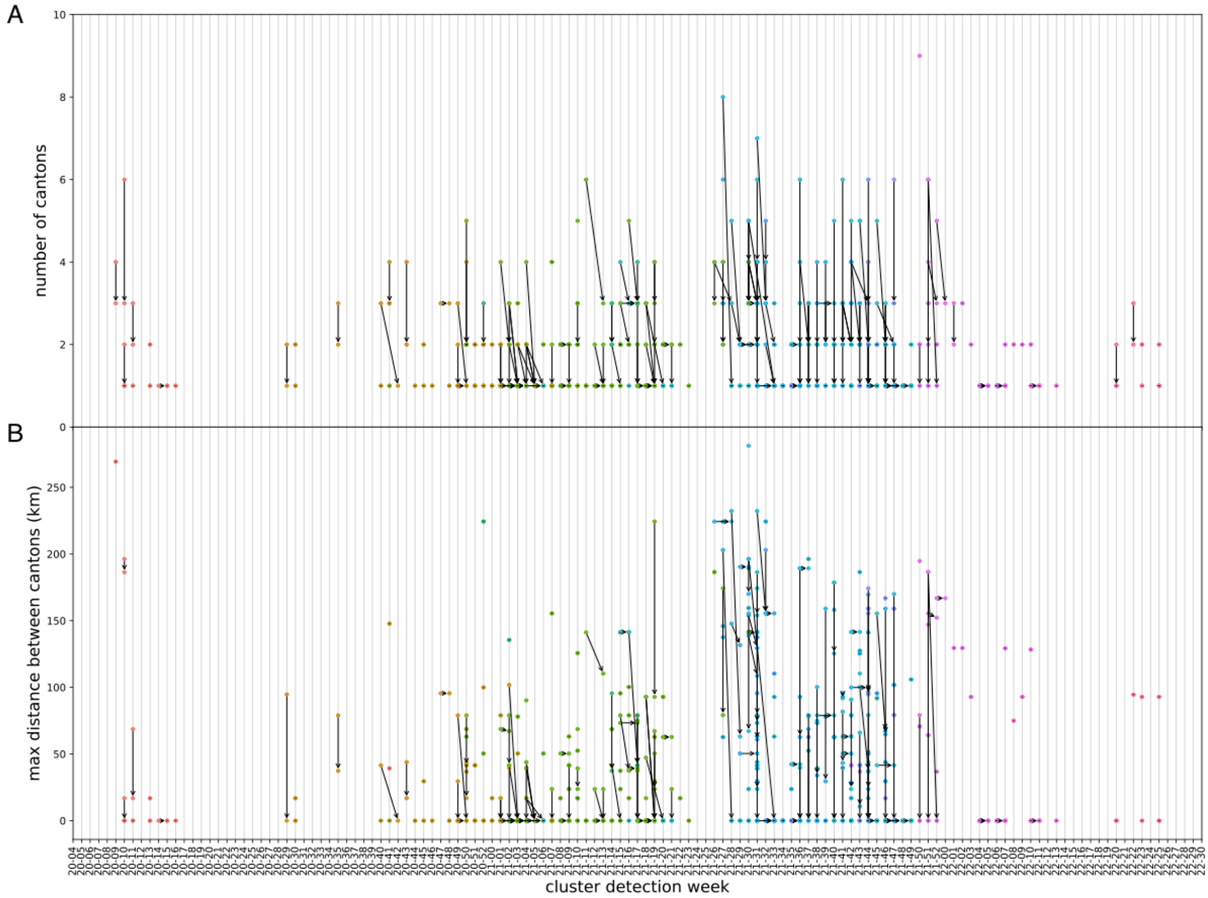


**Figure S9** Shift in geographic spread of clusters after downsampling. (A) Geographic spread represented by the number of cantons affected by a cluster. (B) Geographic spread represented by the maximum distance between the cantons affected by each cluster. The dots represent clusters coloured by VOC (Alpha in green, Delta in blue, and Omicron in pink). Dots connected with an arrow represent the same cluster before and after downsampling (143k to 50k samples). Dots without an arrow indicate the clusters for which no shift occurred. Unsuccessfully matched clusters are not represented. The arrow direction indicates the shift in number of affected cantons and the cluster detection date after downsampling.

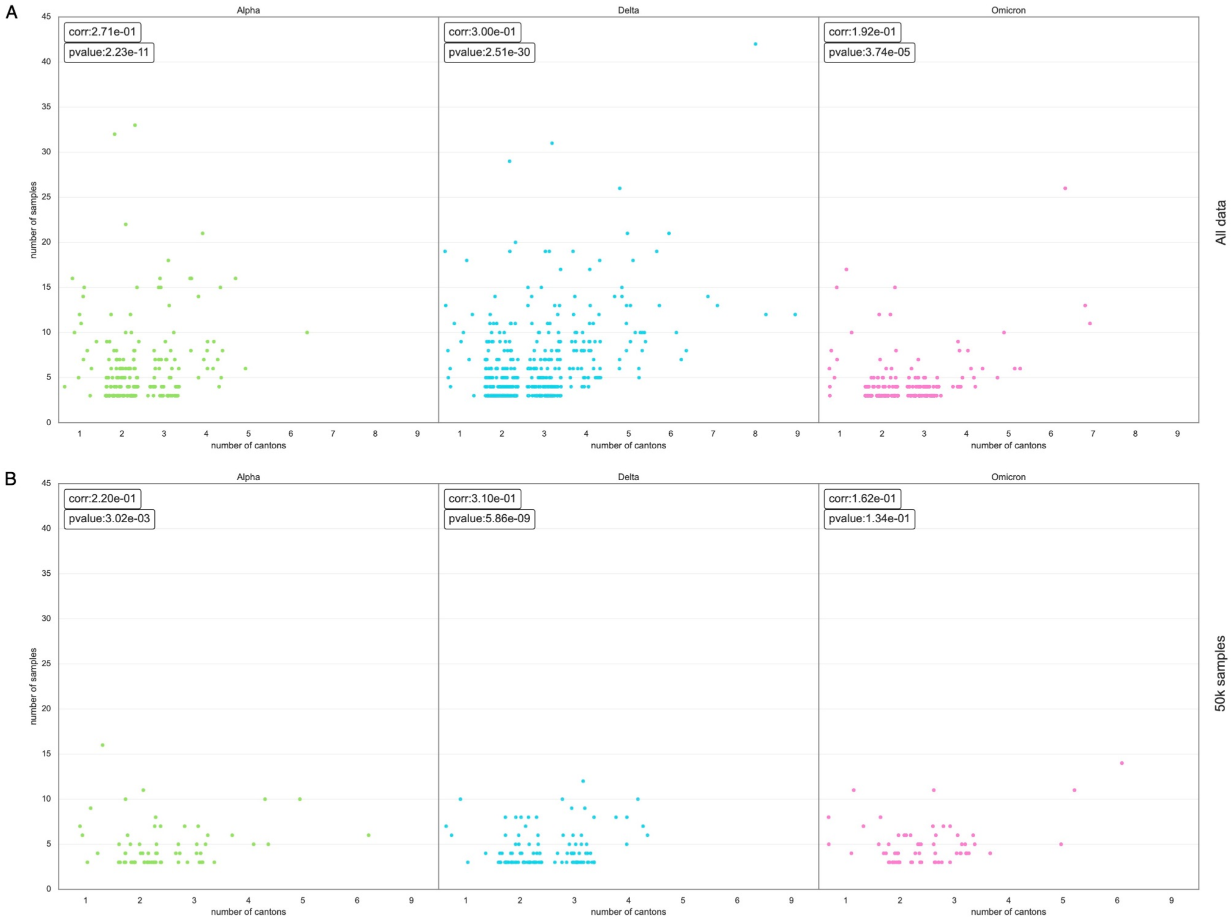


**Figure S10** Correlation between cluster size and number of cantons affected per VOC. We show the correlation between the number of samples in each cluster and the number of cantons affected in the cluster. (A) The results were obtained when considering all the samples in the SPSP dataset. (B)The results obtained when using 50k samples. Each panel represents a VOC.

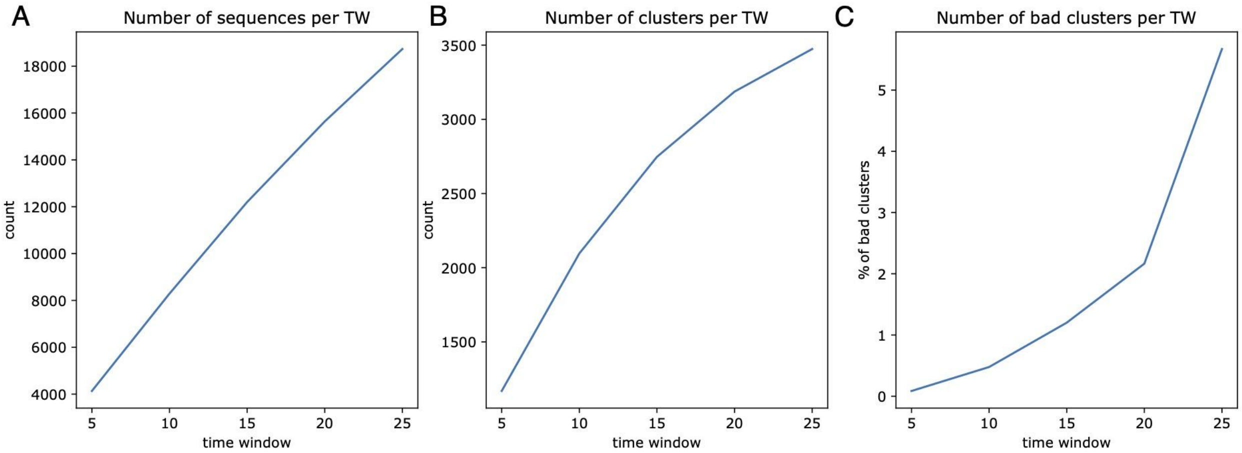


**Figure S11** Analysis of 0-SNP cluster time-windows. (A) Number of sequences analysed per time window. (B) Number of superclusters found for each 0-SNP cluster time window. (C) Number of superclusters spanning more than 30 days for each 0-SNP cluster time window.
